# Spatially resolved mapping of cells associated with human complex traits

**DOI:** 10.1101/2024.10.31.24316538

**Authors:** Liyang Song, Wenhao Chen, Junren Hou, Minmin Guo, Jian Yang

## Abstract

Depicting spatial distributions of disease-relevant cells is crucial for understanding disease pathology. Here, we present a method, gsMap, that integrates spatial transcriptomics (ST) data with genome-wide association study (GWAS) summary statistics to map cells to human complex traits, including diseases, in a spatially resolved manner. Using embryonic ST datasets covering 25 organs, we benchmarked gsMap through simulation and by corroborating known trait-associated cells or regions in various organs. Applying gsMap to brain ST data, we revealed that the spatial distribution of glutamatergic neurons (glu-neurons) associated with schizophrenia more closely resembles that for cognitive traits than that for mood traits, such as depression. The schizophrenia-associated glu-neurons were distributed near the dorsal hippocampus, with upregulated calcium signaling and regulation genes, while the depression-associated glu-neurons were distributed near the deep medial prefrontal cortex, with upregulated neuroplasticity genes. Our study provides a method for spatially resolved mapping trait-associated cells and demonstrates the gain of biological insights (e.g., spatial distribution of trait-relevant cells and related signature genes) through these maps.

## Introduction

The composition and spatial organization of cells within a tissue are critical to its function and can also serve as indicators of its health status^1^. For instance, the radial spatial arrangement of hepatocytes in liver lobules facilitates efficient metabolism of nutrients in the blood^2^, while the spatial distribution and connectivity of neurons in the brain orchestrate emotion, cognition, and behavior^3^. The advancement in spatial transcriptomics (ST), which enables the profiling of gene expression levels of cells in their native spatial positions, represents a promising avenue for investigating cell spatial organizations and uncovering related biological mechanisms^4,5^. In recent years, an increasing number of studies have utilized ST technologies to explore cell spatial organizations in diverse tissues ^6-11^. However, not all cells within tissues are pertinent to a disease or trait of interest. Identifying which cells in a tissue are most relevant to a disease and mapping their spatial distribution is crucial for comprehending disease pathology, yet a substantial knowledge gap remains.

To identify trait-associated cells or cell types, previous studies have proposed genetics-informed strategies that integrate data from genome-wide association studies (GWAS) of complex traits, including diseases, with single-cell RNA sequencing (scRNA-seq) data^12-14^. The fundamental principle behind this integration involves assessing whether genetic association signals for a trait are enriched in or around genes highly expressed in a specific group of cells. Although these methods can pinpoint trait-associated cells, they encounter challenges in mapping the spatial distribution of these identified cells due to the lack of cell spatial positions in scRNA-seq data. These scRNA-seq-based methods can, in principle, be applied to ST data. However, due to the absence of modeling for cell spatial coordinates and the high level of technical noise in ST data^15^, they have limited power in spatially aware mapping of trait-associated cells (see below). Consequently, there is a need for new methods capable of integrating ST data into GWAS for spatially resolved mapping of trait-associated cells.

In this study, we introduce genetically informed spatial mapping of cells for complex traits, gsMap, a method that integrates high-resolution ST data and GWAS summary statistics for spatially resolved mapping of trait-associated cells. Utilizing embryonic ST datasets covering 25 organs, we assessed the specificity of gsMap through simulated GWAS data and the sensitivity of the method by recapitulating known associations between cells in different organs and a range of complex traits. Applying gsMap to brain ST datasets, we generated extensive trait-brain cell association maps encompassing 33 human brain-related complex traits. From these maps, we discovered that spatial patterns of schizophrenia (SCZ)-associated neurons more closely resemble those associated with cognitive traits compared to those associated with mood traits such as depression. Specifically, glu-neurons in the hippocampal cornu ammonis area 1 (CA1) were strongly associated with SCZ, and this association became more pronounced as their spatial distribution progressed from the ventral side to the dorsal side of CA1, with upregulated calcium signaling and regulation genes. Glu-neurons in the gyrus rectus were strongly associated with depression, and this association strengthened as their distribution moved from the lateral side to the medial side of the gyrus rectus, with upregulated neuroplasticity genes. By integrating with drug databases, we found that genes highly expressed in the medial side of gyrus rectus showed a 16.0-fold enrichment in the target genes of psychiatric drugs, demonstrating the clinical value of spatially resolved mapping of disease-associated cells.

## Results

### Overview of gsMap

The fundamental concept of gsMap involves assessing whether genetic variants, predominantly single nucleotide polymorphisms (SNPs), located in or near genes highly expressed in a spot in ST data are enriched for genetic associations with a trait of interest. Here, a ‘spot’ refers to a cell in high-resolution ST platforms (e.g., Stereo-seq cell-bin mode) or a cluster of cells in conventional ST platforms (e.g., 10X Visium). Our gsMap consists of three steps. First, gsMap leverages spot homogeneity to address sparsity and technical noise in ST data. gsMap employs a graph neural network (GNN) to identify homogeneous spots for each focal spot in terms of both gene expression patterns and spatial positions. The gene specificity scores (GSS) of each spot are computed by aggregating information from these homogeneous spots, representing the relative rank of the expression level of each gene in a spot (**Fig. 1a**). Second, gsMap assigns GSS of each spot to SNPs based on their distance from gene transcription start sites (TSS), along with SNP-to-gene maps established using epigenomic data^16-18^, giving rise to a unique set of SNP GSS annotation for each spot. Treating each spot as an SNP annotation set, gsMap assesses whether SNPs with higher GSS disproportionately explain a larger proportion of heritability for the trait using the stratified linkage disequilibrium score regression (S-LDSC)^19,20^, conditional on the baseline annotations (**Fig. 1b**). The enrichment *P* value is used to measure the statistical significance of association of a spot with the trait. Finally, to quantify the significance of association of a specific spatial region with a trait, gsMap employs the Cauchy combination test^21^ to aggregate *P* values of individual spots within that spatial region (**Fig. 1c**). In essence, gsMap can be considered as a method for genetically informed mapping of complex traits to ST data at cellular resolution. We have optimized the computing efficiency of the gsMap to ensure its applicability to large-scale ST data. With prepared annotation files, gsMap took less than 2 hours (one CPU) to map one GWAS trait to ST data with 100K spots. Further details of gsMap can be found in the Methods section.

**Figure 1.**
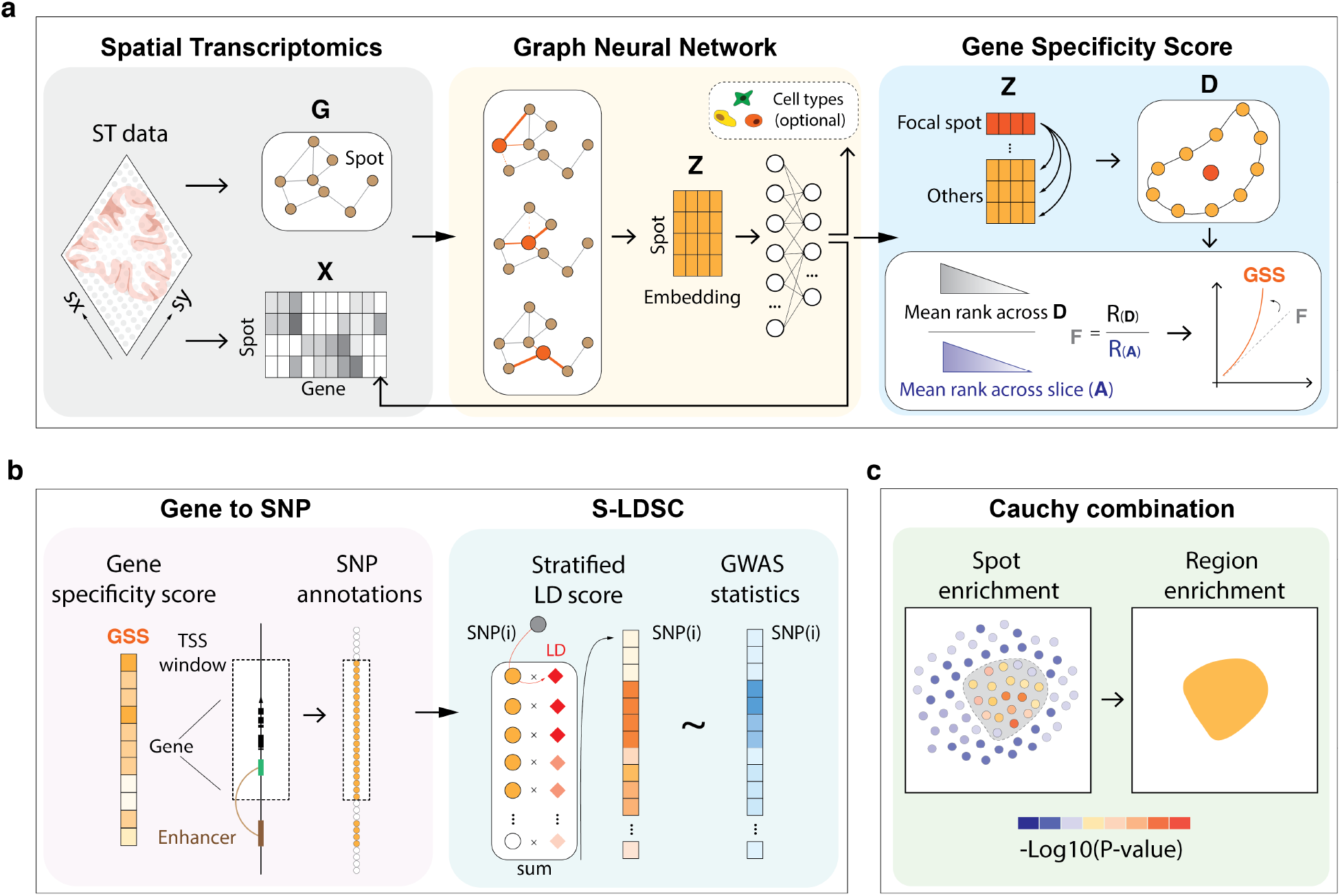
Schematics of the gsMap method. (a) gsMap begins by learning embeddings that integrate gene expression levels, spatial coordinates, and cell type annotation priors through a GNN. Subsequently, gsMap identifies homogeneous spots for each focal spot, based on their cosine similarity in the embeddings, to form a micro-domain. Dividing the rank of a gene’s expression level averaged across all spots in the micro-domain by its average rank across the entire ST data, gsMap computes a specificity score of each gene for each focal spot. (b) The gene specificity scores (GSS) of each spot are then mapped to SNPs, based on their distance to TSS and SNP-to-gene linking maps, resulting in a unique set of SNP annotations for each spot. For the SNP GSS annotations of each spot, gsMap uses S-LDSC to assess whether SNPs with higher GSS are enriched for heritability for the trait of interest. (c) To quantify the significance of a spatial region’s association with the trait, gsMap employs the Cauchy combination test to aggregate *P* values of spots within that spatial region. G: spot spatial graph; X: gene expression matrix; Z: embeddings; D: micro-domain of spots; F: gene expression specificity; GSS: gene expression specificity after Gaussian kernel projection.

### Validation of gsMap

As a proof of principle, we sought to validate gsMap by corroborating known associations between different tissues and traits. For this validation analysis, we used ST data collected from the late embryo stage, as it covers a comprehensive spectrum of tissues, and the gene expression profiles at this stage are similar to those of adult organs. Due to the lack of human embryonic ST data, we used a mouse dataset from the Chen et al. study^6^, which included 25 different organs at the developmental stage of E16.5 (**Fig. 2a**). Similar strategies have been adopted previously, albeit with different mouse datasets^13,22^. This strategy assumes that gene expression profiles in mice resemble those in humans, an assumption largely supported by previous evidence demonstrating over 80% correlation in expression across genes between mice and humans^23^. The Chen et al. mouse ST datasets were generated using the Stereo-seq, containing data at the ‘cell-bin’ resolution, where each spot represents an individual cell, and the ‘bin50’ resolution, where each spot represents a few cells. For robustness, we began by using data at the ‘bin50’ resolution. We included publicly available GWAS summary statistics for 110 complex traits (average N=385K, **Supplementary Table 1**) in this analysis. Our gsMap successfully recapitulated known associations between different organs and traits^12,22^. For example, intelligence (IQ) was mapped to spots located in the brain, mean corpuscular hemoglobin concentration (MCHC) was associated with those in the liver, and height was mapped to various tissues, with cartilage and its primordium showing the highest relevance (**Fig. 2b**). The aggregated tissue-level association *P* values, obtained using the Cauchy-combination test, confirmed these observations (**Fig. 2c**). To further validate the reliability of the identified associations between traits and spots, we simulated null scenarios in which the causal variants were randomly distributed across the genome, thereby not enriched in any gene regions, with varying levels of heritability and polygenicity (Methods). The simulation was based on real genotype data on 100K unrelated individuals of European ancestry from the UK Biobank (UKB)^24^. The false discovery rate of gsMap was well controlled, as it did not identify any spots showing significant associations at FDR<0.05 in the null simulations (**Supplementary Fig. 1**)

**Figure 2.**
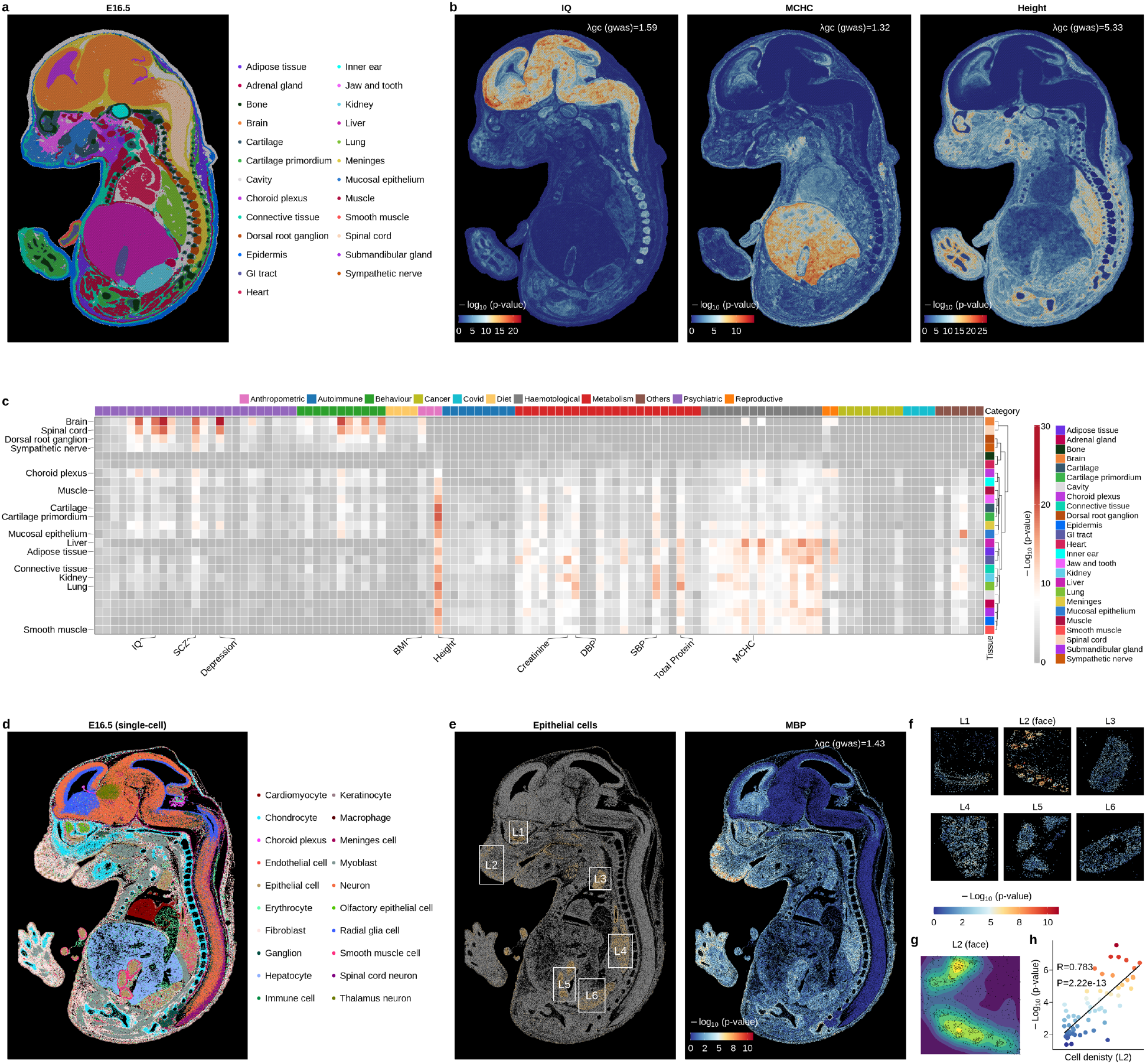
Genetically informed mapping of complex traits to embryonic ST data. (a) Mouse E16.5 embryonic ST data at bin50 resolution, where spots are colored by their tissue types. (b) gsMap results for IQ, MCHC, and height using the mouse E16.5 embryonic ST data, with colors indicating the significance of spot-trait associations. The genomic inflation factor of the GWAS summary statistics, *λ*_GC_, serves as an indicator of the statistical power of the GWAS data. (c) Heatmap of tissue-trait associations obtained using the Cauchy combination test, with colors indicating the significance of the associations and the colored annotations on the right indicating tissue types. Each row corresponds to a tissue type, and each column represents a trait. (d) Mouse E16.5 embryonic ST data at single-cell resolution, where spots are colored by their cell types. (e) Left: Distribution of epithelial cells in the mouse E16.5 embryo, with the white rectangles showing six different spatial regions. The brown dots represent epithelial cells. Right: gsMap results for MPB, where spots are colored according to the significance of their associations with MPB. (f) gsMap results for MPB in the six spatial regions. (g) Densities of epithelial cells in the face (L2) region. The gradient colors from blue to yellow represent cell density from low to high. The black dots represent epithelial cells. (h) Scatter plot showing the correlation between epithelial density and the significance of epithelial cells’ association with MPB in the face region. Epithelial densities were estimated as the smoothed relative count of epithelial cells within 100 grids, while the significance of epithelial-MPB associations for each grid were estimated using the interpolate method. The x-axis shows epithelial density; the y-axis shows the significance of epithelial-MPB association. Each data point represents an individual grid, colored by the significance of its association with MBP. The black line is the regression line.

To further showcase the capability of gsMap in spatially resolved mapping of cells to a trait, we applied gsMap to the embryonic ST data with higher resolution^6^, in which each spot represented an individual cell (**Fig. 2d**). Taking male pattern baldness (MPB) as an example, we identified an association between MPB and epithelial cells in the face region (**Fig. 2e**). Notably, the MPB-associated epithelial cells were not randomly distributed but tended to cluster spatially together (**Fig. 2f**). The significance level of the cells’ association with MPB was strongly correlated (*r* = 0.78 and *P*=2.2e-13) with the density of epithelial cells (**Fig. 2g-h**). The tight spatial assembly of epithelial cells suggested that they may form hair follicles^25^, supported by the expression profile of hair follicle-associated marker genes – *KRT15, KRT5*, and *KRT17* (**Supplementary Fig. 2**).

As mentioned above, established methods for integrating GWAS with scRNA-seq data, such as scDRS^13^, can be repurposed for ST data, even though spatial coordinate information is not accounted for in their models. Hence, we performed a comparison between gsMap and scDRS using both simulated and real GWAS datasets. We first evaluated the methods with the simulated GWAS data under the null and the mouse E16.5 embryonic ST data at the bin50 resolution. As aforementioned, the false discovery rate of gsMap was well controlled, without identifying any significant spots (0/121,335) at FDR<0.05 under the null, across 12 different simulation settings and 3 independent replicates (**Supplementary Fig. 1**). However, scDRS showed an average of 2,857 false-positive spots (2,857/121,335) at FDR<0.05 across simulation settings and replicates, and the false discovery rate increased with the increasing statistical power of the GWAS data (**Supplementary Fig. 3**). Next, to compare the power of gsMap and scDRS in real data analysis, we curated well-established trait-tissue association pairs, including IQ with the brain, low-density lipoprotein (LDL) with the liver, and creatinine with the kidney. gsMap clearly showed the association of IQ with the brain (*P*=1.9e-19), LDL with the liver (*P*=2.8e-8), and creatinine with the kidney (*P*=3.3e-14), while the corresponding *P* values from scDRS – 0.0012 (IQ-brain), 0.47 (LDL-liver), and 0.25 (creatinine-kidney) – were not significant after correcting for multiple testing (**Supplementary Fig. 4**). The scDRS results for all traits are available at gsMap data portal: https://yanglab.westlake.edu.cn/gsmap/home.

### Genetically informed mapping of human complex traits to brain regions at cellular resolution

We first applied gsMap to map human complex traits to brain regions, largely due to the abundant availability of brain ST data, even though the majority is not derived from human samples. To ensure statistical power, we included 33 distinct brain-related traits with large GWAS sample sizes (average N=410K) in our analyses, which cover cognition, emotion, and behavior. Due to the lack of human whole brain ST data, we began by integrating GWAS summary statistics with mouse brain ST data and primarily focused on the evolutionarily conserved brain regions to ensure the broad extrapolation of the results. We processed adult mouse hemibrain coronal section ST data from the Chen et al. study^6^, which comprised 50,140 cells from 14 brain regions and 13 cell types (**Fig. 3a**). We started with assessing the association of an entire brain region with a trait by aggregating the *P* values of individual cells in each brain region using the Cauchy combination test (**Fig. 3b**). We observed that the cortex showed the highest relevance to most traits, such as IQ (*P*=9.8e-15), SCZ (*P*=1.7e-17), and depression (*P*=1.0e-18). The second-highest associated brain regions varied across traits: IQ was mapped to the hippocampus CA1 (*P*=7.2e-13), SCZ to the hippocampus CA1 (*P*=4.4e-14), and depression to the midbrain (*P*=1.1e-15).

**Figure 3.**
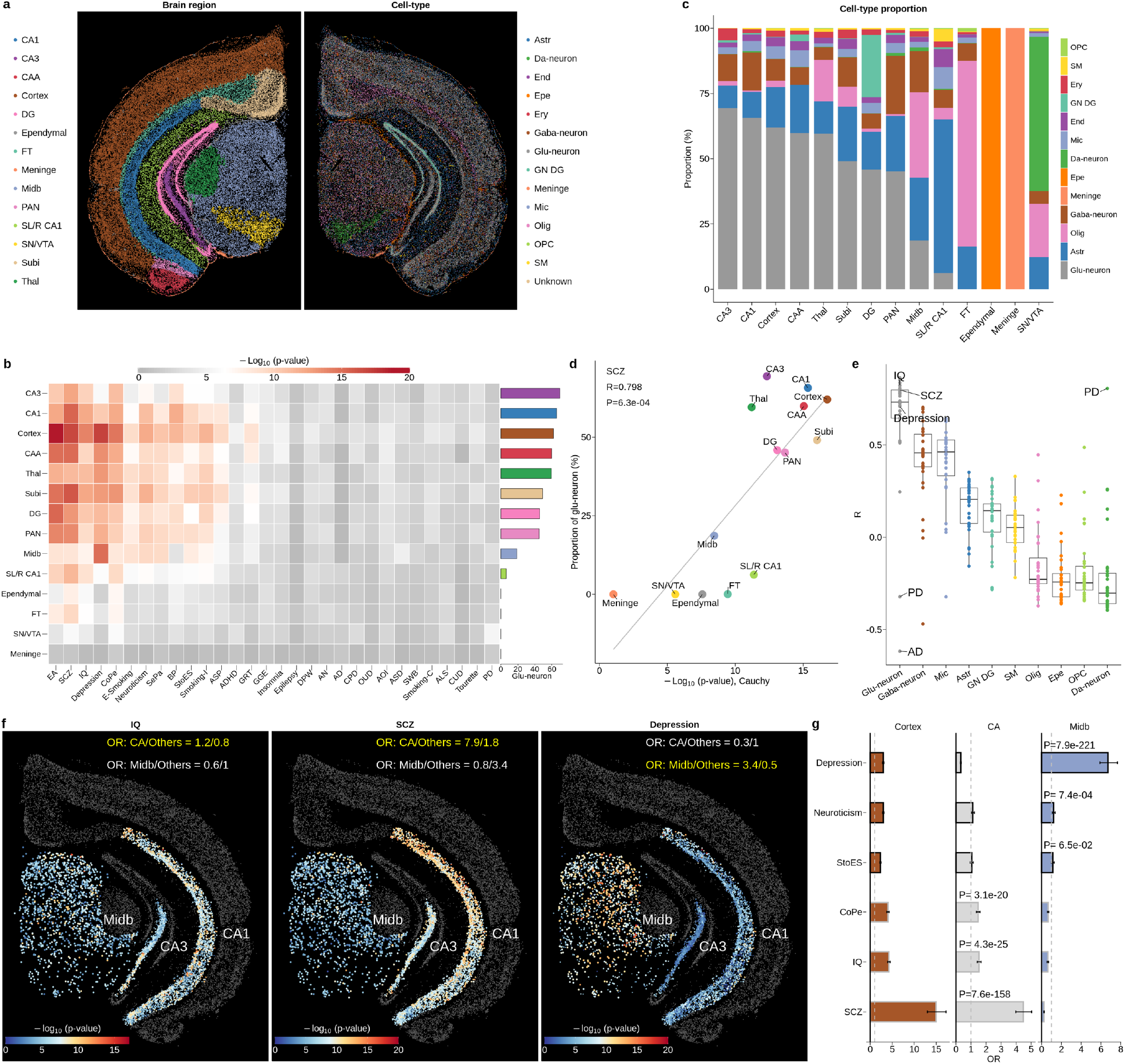
Genetically informed mapping of complex traits to adult mouse brain ST data. (a) Adult mouse brain ST data, where spots are colored by their brain regions (Left) or cell types (Right). CA1, cornu ammonis area 1; CA3, cornu ammonis area 3; CAA, cortical amygdala area; DG, dentate gyrus; FT, fiber tract; Midb, midbrain; PAN, posterior amygdala nucleus; SL/R CA1, stratum lacunosum/raditum cornu ammonis area 1; SN/VTA, substantia nigra/ventral tegmental area; Subi, subiculum; Thal, thalamus. Astr, astrocyte; Da-neuron, dopaminergic neuron; Endo, endothelial cell; Epe: ependymal; Ery, erythrocyte; Gaba-neuron, GABAergic neuron; Glu-neuron, glutamatergic neuron; Mic, microglia; Oligo, oligodendrocyte; OPC, oligodendrocyte precursor cell; SMC, smooth muscle cell. (b) Heatmap of brain region-trait associations obtained using the Cauchy combination test, with colors indicating the significance of the associations and the colored bar on the right representing the proportion of glu-neurons in each brain region. Each row corresponds to a brain region, and each column represents a trait. (c) Bar plot displaying cell type proportions in each brain region. (d) Correlation between the significance of a brain region’s association with SCZ (x-axis) and its proportion of glu-neurons (y-axis). The grey line represents the regression line. (e) Boxplot showing the correlation between the significance of association with a trait and the proportion of a cell type across brain regions. The x-axis displays different cell types; the y-axis shows the Pearson correlation coefficient. Each point represents a trait, with its color indicating the corresponding cell type. (f) gsMap results for IQ, SCZ, and depression in the midbrain and CA areas. Each point represents an individual glu-neuron, with color indicating the significance of its association with a trait. (g) Bar plots displaying the OR of Cortex (Left), CA (Middle), and Midb (Right), respectively. Each error bar represents the 95% confidence interval (CI) of an OR. StoEs, sensitivity to environmental stress; CoPe, cognitive performance.

As brain regions contain various cell types, we explored their contributions to the brain region-trait associations by assessing whether a higher proportion of a specific cell type correlates with increased significance in brain region-trait association. We discovered that, on average across traits, glu-neurons showed the strongest contributions to the brain region-trait associations (median *r* = 0.73) among all cell types. For instance, the correlation between the significance of a brain region’s association with SCZ and its glu-neuron proportion was 0.80 (*P*=6.3e-04) (**Fig. 3c-e**). Such a relationship was replicated using brain sagittal section ST data from the mouse E16.5 embryo (**Supplementary Fig. 5**). However, there were two outliers, Parkinson’s Disease (PD) and Alzheimer’s Disease (AD). We observed that brain regions with higher proportions of dopaminergic neurons (da-neurons) exhibited stronger associations with PD (*r* = 0.81 and *P*=4.9e-4; **Fig. 3e**). Specifically, the substantia nigra/ventral tegmental area (SN/VTA), known for housing the most abundant da-neurons, demonstrated the highest relevance to PD (*P*=5.2e-5; **Supplementary Fig. 11**). This finding was in line with previous neurological studies that showed a notable impairment in the SN/VTA region among PD patients^26,27^.

Having noted the important role of glu-neurons in most of the analyzed traits, we proceeded to explore whether the associated glu-neurons were distributed in distinct patterns for different traits. Recognizing that direct comparison of gsMap results across traits might be confounded by variation in GWAS statistical power, we employed odds ratio (OR), calculated as the ratio of trait-associated glu-neurons to non-associated ones in a specific region, divided by that ratio in all other regions (Methods). Our analysis focused on SCZ, cognitive traits (e.g., IQ), and mood traits (e.g., depression), as glu-neurons significantly contributed to these traits’ associations with brain regions (**Fig. 3e**), and their GWAS data were sufficiently powered (*λ*_GC_>1.3). We found that, beyond the cortex, glu-neurons in the hippocampus CA areas were strongly associated with cognitive traits (e.g., *OR*=1.5 and *P*=4.3e-25 for IQ), while glu-neurons associated with mood traits tended to be distributed in the midbrain (e.g., *OR*=6.7 and *P*=7.9e-221 for depression; **Fig. 3f-g**). Interestingly, although SCZ encompasses both cognitive and mood symptoms, we found that glu-neurons associated with SCZ exhibited a similar spatial pattern to cognitive traits, particularly enriched in the hippocampus CA areas (*OR*=4.5 and *P*=7.6e-158; **Fig. 3g**).

Taken together, our results revealed that: 1) among the brain regions, the cortex exhibited the strongest association with most traits, but PD was only significantly associated with the SN/VTA; 2) glu-neurons substantially contributed to the associations between traits and brain regions; 3) glu-neurons distributed in different brain regions displayed varied associations with traits. Glu-neurons distributed in the hippocampus CA areas showed stronger associations with cognitive traits and SCZ, while those in the midbrain exhibited stronger associations with mood disorders such as depression.

### Spatial distributions of trait-associated glu-neurons in the hippocampus

Having shown that the ratio of trait-associated glu-neurons varied in different brain regions, we next sought to explore whether the trait-associated glu-neurons exhibit a specific pattern of spatial distribution within a brain region. We began by focusing on the hippocampus cornu ammonis area 1 (CA1), as the spatial structure of the CA1 region is relatively simple, and previous studies^28-30^ have suggested that the electrophysiological properties of pyramidal neurons in CA1 exhibit variation across their spatial positions. Cells within the CA1 region are densely arranged along three spatial axes: the proximal-distal (P-D) axis, the dorsal-ventral (D-V) axis, and the deep-superficial (D-S) axis (**Fig. 4a**). Since the brain ST data used in this study correspond to a coronal section, tracking the spatial distribution of glu-neurons along the P-D axis was unfeasible. Consequently, our investigation focused on the spatial distribution of glu-neurons along the D-V and D-S axes.

**Figure 4.**
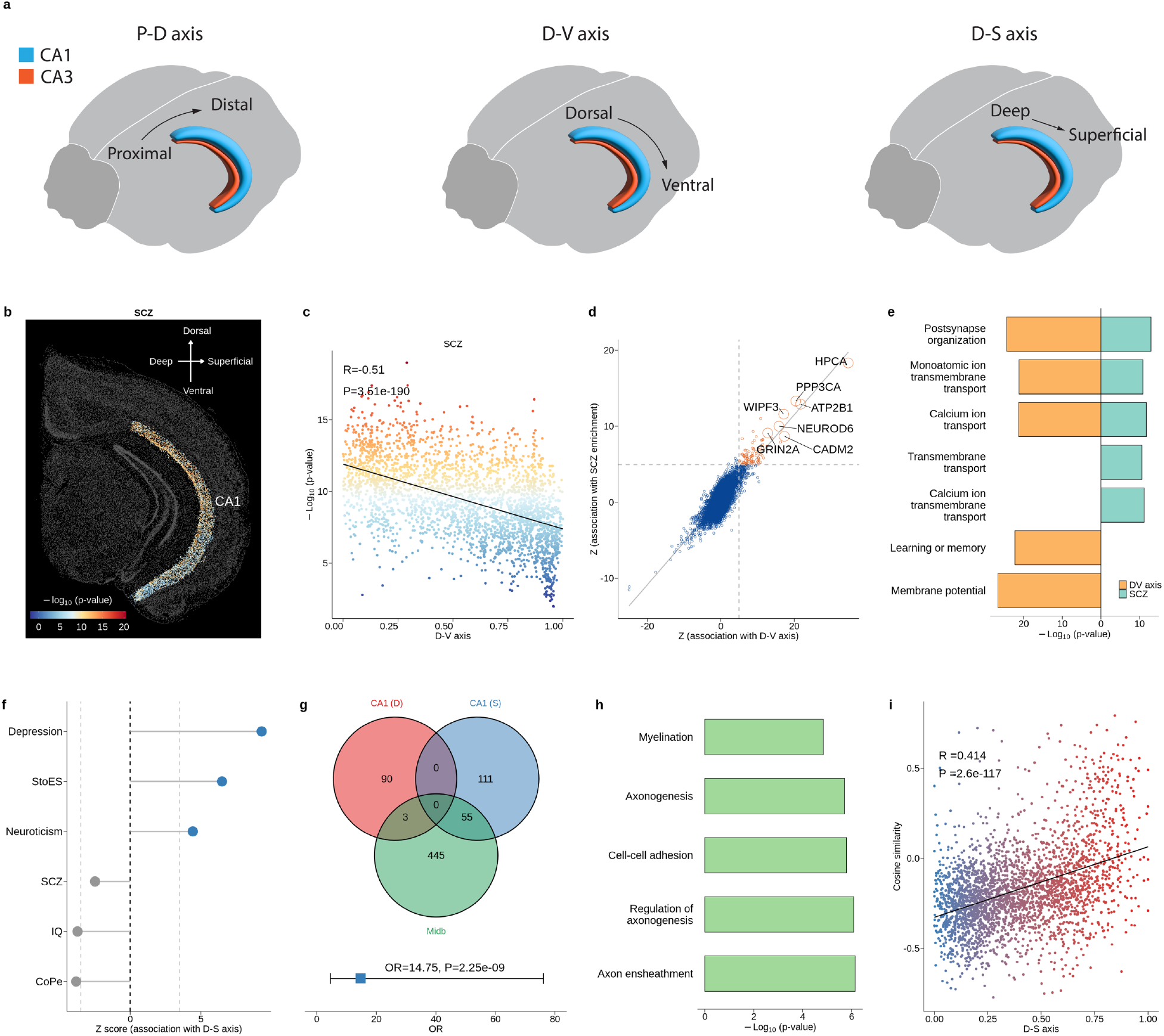
Spatial distributions of trait-associated glu-neurons in the hippocampus. (a) Schematics of the three spatial axes of the mouse hippocampus CA1/CA3 region. (b) gsMap results of SCZ in mouse brain ST data, with colors indicating the significance of associations of cells in the hippocampus CA1 region with SCZ. (c) Correlation between the significance of a glu-neuron’s association with SCZ and its spatial position along the CA1 D-V axis. Each data point represents an individual glu-neuron in the CA1 region, with the color indicating the significance of its association with SCZ. The black line denotes the regression line. (d) Scatter plot of Z-statistics from the DEG analyses. The x-axis shows the Z-statistic for correlation between the expression levels of a gene in glu-neurons and their spatial positions along the D-V axis; the y-axis shows the Z-statistic for correlation between the gene expression levels in glu-neurons and the significance of their associations with SCZ. Each data point represents one gene, and those with FDR<0.01 are highlighted in orange. (e) GO enrichment analysis of genes whose expression levels were significantly correlated with glu-neurons’ spatial positions along the D-V axis (orange) or the significance of glu-neurons’ associations with SCZ (cyan). (f) Z-statistic for the correlation between the significances of glu-neurons’ association with a trait and their spatial positions along the CA1 D-S axis. Each data point represents a specific trait, and the grey dashed line represents the significant threshold at FDR=0.01. (g) Top: Venn diagram showing the overlap among genes highly expressed in the CA1 deep side, the CA1 superficial side, and the midbrain; Bottom: odds ratio (OR) contrasting the number of genes highly expressed in both the midbrain and CA1 superficial side to that highly expressed in both the midbrain and CA1 deep side. The blue square represents the OR, with the error bar showing the 95% CI. (h) GO enrichment analysis of genes highly expressed in both the midbrain and CA1 superficial side. (i) Correlation between the spatial positions of CA1 glu-neurons along the D-S axis (x-axis) and the cosine similarity of gene expression levels between glu-neurons in the CA1 and in the midbrain (y-axis). Each data point represents an individual glu-neuron, colored according to its position along the D-S axis. The black line denotes the regression line.

First, along the D-V axis, we observed that glu-neurons closer to the CA1 dorsal side showed stronger associations with the traits (**Fig. 4b** and **Supplementary Fig. 7**), with SCZ showing the strongest association (*r* = −0.51 and *P*=3.5e-190; **Fig. 4c**). To investigate the underlying mechanisms, we conducted a differential gene expression (DEG) analysis to identify genes whose expression levels in glu-neurons were correlated with their positions along the D-V axis and a DEG analysis to identify genes in glu-neurons whose expression levels were correlated with their relevance to SCZ. The *Z* statistics between these two DEG analyses showed a strong correlation (*r* = 0.78 and *P*<5e-324), with *HPCA* (encoding a calcium-binding protein), *PPP3CA* (encoding a calcineurin subunit), and *ATP2B1* (encoding a calcium-transporting protein) emerging as the top three genes prioritized in both DEG analyses (**Fig. 4d**). A subsequent gene ontology (GO) enrichment analysis suggested that the identified genes were enriched in pathways related to calcium transport and ion transport (**Fig. 4e**).

The spatial scale of the D-S axis is narrow, yet we also observed heterogeneity among glu-neurons along this axis. Leveraging gsMap, we found that glu-neurons closer to the CA1 superficial side exhibited stronger associations with depression (*r* = 0.18 and *P*=2.7e-20; **Fig. 4f** and **Supplementary Fig. 8**). This finding aligned with a recent study suggesting that manipulating CA1 superficial neurons, but not deep neurons, could ameliorate depressive-like behaviors in mice^31^. Motivated by the result above that glu-neurons in the midbrain were highly relevant to depression (**Fig. 3g**), we further investigated associations between glu-neurons in the CA1 superficial side and those in the midbrain. We found that genes highly expressed (at FDR<0.05) in the midbrain showed a greater overlap with genes highly expressed in the CA1 superficial side than those in the CA1 deep side (55 vs. 3 genes, *OR*=14.75, *P*=2.25e-09, **Fig. 4g**). GO enrichment analysis indicated that genes highly expressed in both midbrain and the CA1 superficial side were enriched in pathways related to axon genesis and neuron ensheathment (**Fig. 4h**). The transcriptomic similarities of glu-neurons between the CA1 and the midbrain increased along the CA1 D-S axis (*r* = 0.41 and *P*=2.6e-117; **Fig. 4i**). These results suggested that there are functional overlaps between glu-neurons in the midbrain and those at the CA1 superficial side.

In summary, our results revealed spatially patterned distributions of trait-associated glu-neurons in the hippocampus CA1 region. Glu-neurons closer to the CA1 dorsal side showed stronger associations with SCZ, featured with increased expression of calcium signaling and regulation genes. Glu-neurons closer to the CA1 superficial side exhibited stronger associations with depression, featured with increased expression of axon genesis-related genes.

### Genetically informed spatial mapping of complex traits to primate cerebral cortex

The hippocampus and midbrain are evolutionarily conserved across different mammalian species in terms of their spatial structures and biological functions^32-34^. Utilizing mouse ST data, we have identified neurons associated with complex traits in the hippocampus and midbrain and revealed that these associations correlated with the spatial distributions of neurons. Next, we sought to extend our analyses to an evolutionarily more advanced brain area—cerebral cortex^35^. Considering the functional and structural differences between the human and mouse cortex^35-37^, conducting a fine-resolution mapping of human traits to mouse cortex ST data presents a challenge. Ideally, human cortex ST data should be utilized. However, the currently available human dataset^9^ is limited by its cortex coverage, which only encompasses a small portion of the dorsolateral prefrontal cortex (DLPFC), and by its resolution, as it was obtained from the 10X Visium platform where one spot represents several dozen cells. Consequently, we opted to utilize the macaque cerebral cortex ST dataset from the Chen et al. study^8^. This dataset more closely resembles the human cortex than the mouse cortex ST dataset^38^, covering 143 cortical regions spanning from anterior to posterior across the entire left cerebral cortex, all at single-cell resolution (**Supplementary Tables 2-3**).

The serial cortex sections of the macaque ST data offer an additional opportunity to validate gsMap, as the adjacent ST sections can be viewed as technical replicates (**Fig. 5a**). To compare the results in different ST sections, we aggregated the association *P* values of individual cells into cell types within a cortical region. Subsequently, we calculated the Pearson correlation (*r*) of –log10(*P* values) between technical replicates across different cell types and cortical regions. The gsMap results showed remarkable consistency across technical replicates (e.g., median *r* = 0.92 for SCZ and 0.90 for depression; **Fig. 5b** and **Supplementary Figs. 9-10**). In addition, while single-cell resolution human whole cortex data were unavailable, we examined the consistency between gsMap results in human DLPFC and macaque prefrontal cortex (PFC). We chose to use the macaque PFC due to the ambiguity in the sampled positions of the human DLPFC, which made it difficult to accurately select the matched cortical lobe from the macaque (Methods). The gsMap results based on the macaque PFC ST data (Stereo-seq) were strongly correlated (*r* = 0.51, *P*=8.6e-8) with those based on the human DLPFC ST data (10X Visium), despite the discrepancies including differences in ST platform and definition of cortical lobe (**Supplementary Fig. 11-12**). Motivated by these results, we mapped 33 human brain-related traits to ST sections across the entire macaque left cerebral cortex at a spatially resolved single-cell resolution. These mapping results are available in our interactive website at https://yanglab.westlake.edu.cn/gsmap/home.

**Figure 5.**
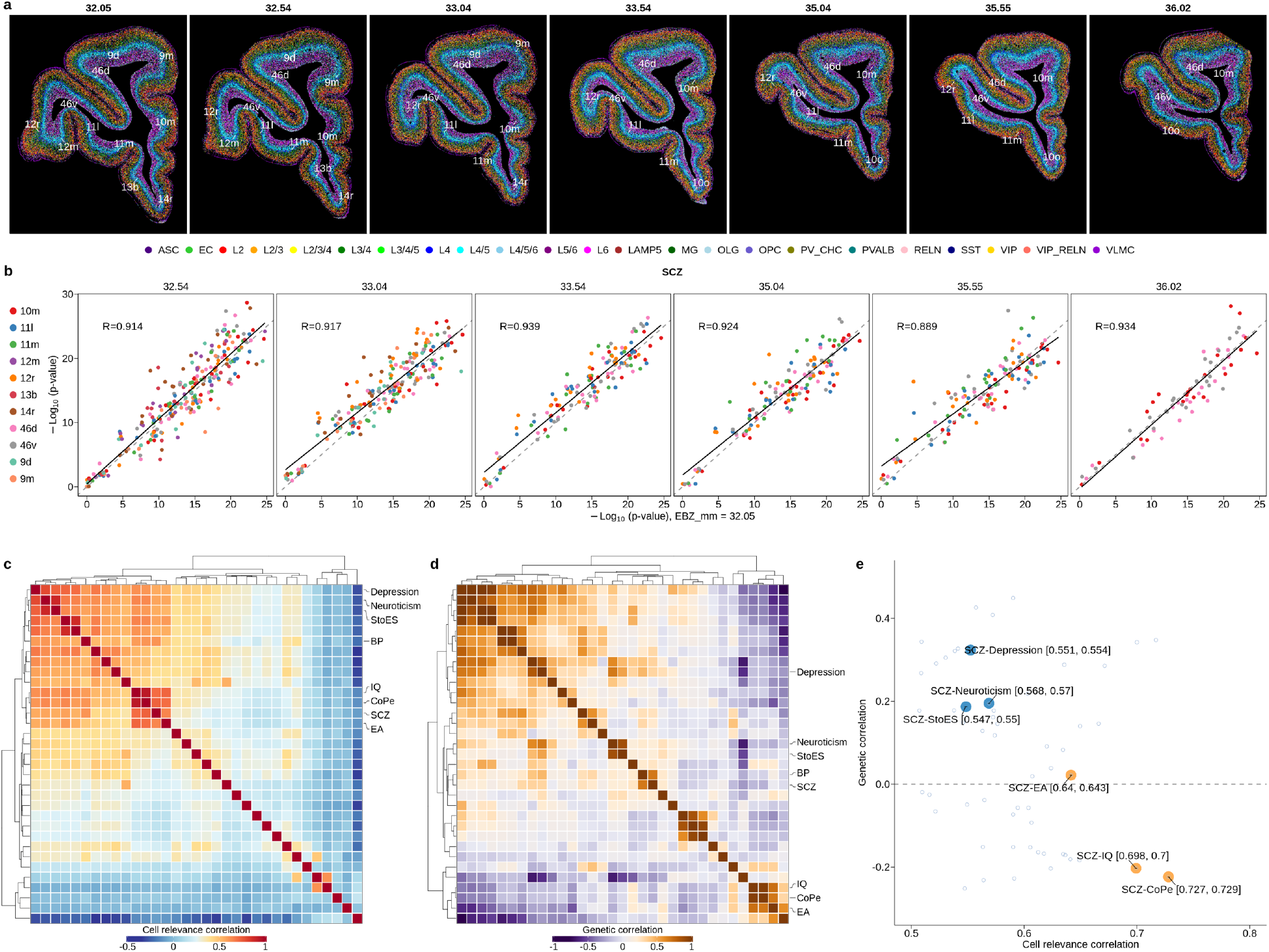
Genetically informed mapping of complex traits to adult macaque cerebral cortex. (a) Representative ST sections of the adult macaque left prefrontal cortex, where cells are colored by cell types. The title of each image displays the cutting position of each ST section, ranging from ear bar zero (EBZ, mm) 32.05 to 36.02. The white texts on the image annotate the cortex region. (b) Cell type-SCZ associations in each of the cortex region estimated using the EBZ 32.05 ST section vs. those estimated using the other ST sections. In each cortex region, we aggregated the individual cells’ associations with SCZ into the association of each cell type using the Cauchy combination test. The x-axis displays the associations of cell types with SCZ estimated using the EBZ 32.05 ST section; the y-axis shows the corresponding associations estimated using the ST sections from EBZ 32.54 to EBZ 36.02. Each data point represents a cell type, colored according to its cortex region. The black line is the regression line, and the grey dashed line represents the line of equality. (c) Heatmap of CRCs between traits estimated using gsMap with the macaque cortex ST data. (d) Heatmap of genetic correlations between traits estimated using the bivariate LDSC method. Each column or row represent a trait. (e) Scatter plot showing the CRCs between traits (x-axis), and the corresponding genetic correlations (y-axis). The orange and blue points highlight the CRCs between SCZ and cognitive-related and mood-related traits, respectively. The texts in square brackets show the 95% CI of CRCs. EA, educational attainment; BP, bipolar disorder.

To systematically analyze these spatial association maps between brain-related traits and cortex cells, we began by estimating the correlation between a pair of traits regarding their associations with cells across the cerebral cortex (referred to as cell relevance correlation or CRC hereafter). We observed that SCZ clustered with other cognitive traits (e.g., cognitive performance, CoPe), while mood traits formed another cluster (**Fig. 5c**). This result was consistent with our earlier discovery using the mouse brain ST data (**Figs. 3**-**4** and **Supplementary Fig. 13**), indicating differences in relevant brain regions between cognitive and mood traits. Notably, CRC between two traits could differ substantially from their genetic correlation (**Fig. 5d**). For instance, while the genetic correlation between glycated hemoglobin (HbA1c) and high-density lipoprotein (HDL) was negative (*r*_*G*_ = −0.18 and *P*=1.6e-11), their CRC was positive (CRC=0.78 and *P*<5.0e-324), due to their shared associations with cells distributed in the liver (**Supplementary Fig. 14**). SCZ is a complex psychiatric disorder that involves a combination of cognitive, mood, and behavioral symptoms, which is typically not classified solely as a cognitive or mood disorder. However, in our study, we found a stronger CRC of SCZ with cognitive traits (e.g., CRC = 0.72 between CoPe and SCZ) across cortex cells (**Fig. 5e**) compared to mood traits (e.g., CRC = 0.55 between depression and SCZ). This result indicated widely shared cells and cortical regions between SCZ and cognitive performance, suggesting that impairment in the brain areas associated with cognition might be a major pathological change in SCZ patients.

### Spatial distributions of trait-associated glu-neurons in the PFC

Leveraging the spatially resolved cortex cell-trait association maps generated above, we investigated the spatial distributions of trait-associated neurons in the adult macaque PFC, considering that previous studies have indicated significant roles of the PFC in emotion and cognition^39-43^ (**Fig. 6a**). We hasten to state that neurons in the other cortex lobes are also crucial, however, due to space limitations, we focused on the PFC lobe here and presented results of other cortex lobes in our interactive website.

**Figure 6.**
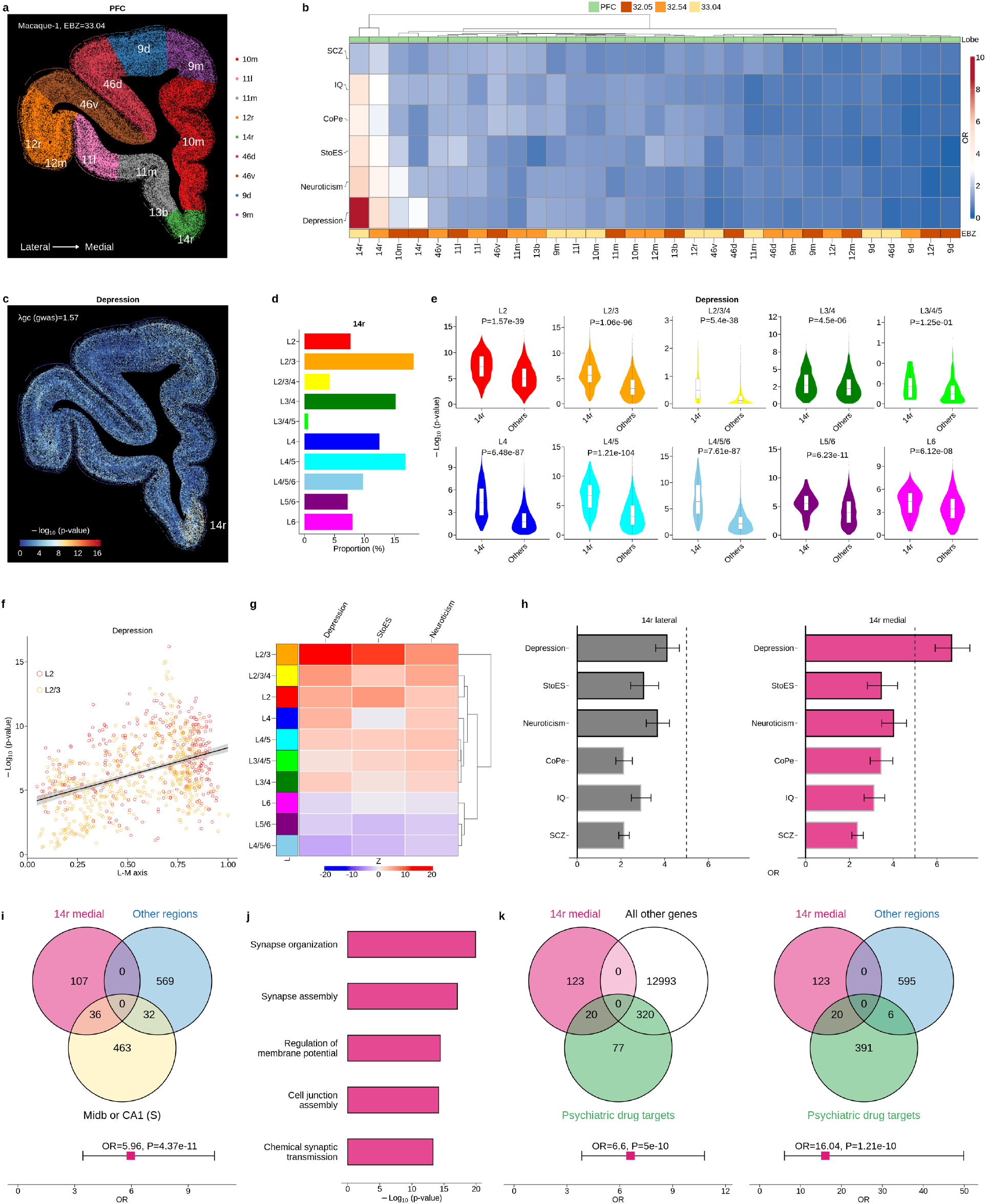
Spatial distributions of trait-associated glu-neurons in the PFC. (a) Adult macaque PFC ST data, where cells are colored by their cortex regions. (b) Heatmap of ORs of glu-neurons (i.e., ratio of trait-associated glu-neurons to the non-associated ones in the focal region divided by that in all other regions), with the colored annotations at the bottom indicating ST sections. Each column corresponds to a cortex region, and each row represents a trait. (c) gsMap results for depression. Each point represents an individual cell, colored by the significance of its association with depression. (d) Bar plot of proportions of glu-neuron subtypes in the 14r region. (e) Boxplot of gsMap results for different glu-neuron subtypes in the PFC 14r region and the other PFC regions, with the text showing the Wilcoxon rank sum test *P* value for the difference. (f) Correlation between the spatial positions of L2 (or L2/3) glu-neurons along the L-M axis (x-axis) and the significance of their associations with depression (y-axis). Each point denotes an individual glu-neuron, colored by its subtype. The black line is the regression line, with the shaded area indicating the 95% CI. (g) Heatmap of Z-scores for correlation between the significance of glu-neurons’ associations with a trait and their positions along the L-M axis. Each row corresponds to a glu-neuron subtype, and each column represents a trait. (h) Bar plot of ORs of glu-neurons in the 14r lateral side (Left) and in the 14r medial side (right), with error bars representing the 95% CIs. (i) Fold-enrichment of genes highly expressed (at FDR<0.05) in the mouse midbrain or CA1 (S) and in the macaque PFC 14r medial side, compared to those highly expressed in the other PFC regions. The red square denotes the OR, with the error bar showing the 95% CI. CA1 (S): hippocampus CA1 superficial side. (j) GO enrichment analysis of genes highly expressed in the macaque PFC 14r medial side. (k) Fold-enrichment of genes highly expressed in the PFC 14r medial side in psychiatric drug targets, compared to all other detected genes (Left) or genes highly expressed in the other PFC regions (Right). The red square denotes the OR, with the error bar showing the 95% CI.

We first explored the distributions of trait-associated glu-neurons in different PFC regions. We discovered that glu-neurons in the PFC 14r region (i.e., gyrus rectus) exhibited a strong association with depression (*OR*=5.3, *P*=5.2e-123, **Fig. 6b-c**), and these results remained consistent when using data from adjacent PFC ST sections (**Supplementary Fig. 15**). This robust association was evident across most subtypes (layers) of glu-neurons, except for rare glu-neurons annotated as L3/4/5 (**Fig. 6d-e** and **Supplementary Fig. 16**). Furthermore, we discovered that the associations between traits and glu-neurons in the PFC 14r region correlated with their local spatial distributions. Along the lateral-medial (L-M) axis of the 14r region, we noticed an increasing relevance of glu-neurons to mood disorders, particularly within the superficial cortex layers (e.g., L2 and L3, **Fig. 6f-g**). When dividing the 14r region into lateral and medial sides, depression (lateral side *OR*=4.1, *P*=2.9e-78; medial side *OR*=6.7, *P*=3.6e-169) exhibited more pronounced associations with glu-neurons in the medial side (**Fig. 6h**). To investigate the underlying biological mechanisms, we conducted a DEG analysis by comparing gene expression levels of glu-neurons in the medial side of the PFC 14r region to those in the other PFC regions. We identified 143 highly expressed genes (at FDR<0.05) in the 14r medial side (**Supplementary Fig. 17**). GO enrichment analysis revealed that these genes were enriched in pathways related to synapse organization and cell junction assembly (**Fig. 6j**). Recalling the findings above that glu-neurons distributed in both the midbrain and CA1 superficial side showed high relevance to depression (**Figs. 3-4**), we also assessed the overlap between these genes and the genes highly expressed in midbrain or CA1 superficial side in the mouse brain ST data. This overlap was significantly higher compared to the overlap with genes highly expressed in the other PFC regions (*OR*=6.0, *P*=4.4e-11, **Fig. 6i**). Moreover, genes highly expressed in the midbrain and CA1 superficial side were enriched in pathways of axon genesis and cell adhesion (**Fig. 4i**), which are closely interconnected with pathways of synapse organization and cell junction assembly identified in the PFC 14r medial side. These pathways collectively contribute to neural plasticity, a fundamental mechanism of neuronal adaptation to environmental stimuli, which might be disrupted in depression patients^44-50^.

Next, to demonstrate the clinical value of the PFC 14r region prioritized by gsMap, we conducted an enrichment analysis to explore whether genes highly expressed in the PFC 14r region are enriched in target genes of approved or launched psychiatric drugs. We collected 417 psychiatric drug target genes from the Drug Repurposing Hub and DrugBank database (Methods)^51,52^. Genes highly expressed in the PFC 14r region showed significant enrichment in psychiatric drug targets, compared to all other genes captured in the macaque ST data (*OR*=5.1, *P*=3.7e-4) or genes highly expressed in the other PFC regions (*OR*=5.2, *P*=1.6e-10; **Supplementary Fig. 18**). This enrichment was even more pronounced for genes highly expressed in the medial side of the PFC 14r region (*OR*=16.0, *P*=2.2e-10; **Fig. 6k**). To validate this result, we estimated the drug module score of individual cells in the PFC ST data, calculated as the average expression level of 417 psychiatric drug target genes minus the average expression level of randomly selected control genes with matched expression characteristics of drug target genes^53,54^ (Methods). Consistently, we observed that cells distributed in the PFC 14r medial side exhibited the highest drug module score compared to those distributed in other PFC regions (*P*=4.2e-27, **Supplementary Fig. 19**). Inspired by these results, we proposed to search for drugs (chemical compounds) whose target genes were enriched in genes highly expressed in the PFC 14r medial side compared to all other genes. The *P* value was determined using the Fisher exact test. We examined all (76,311) drugs/chemical compounds listed in the aforementioned drug databases but only computed OR and *P* values for drugs containing at least 3 known targets. We identified 112 drugs/chemical compounds, including acamprosate (*OR*=32.3, *P*=1.5e-7), pentobarbital (*OR*=27.7, *P*=3.2e-7), gabapentin (*OR*=18.5, *P*=1.7e-5), and others (**Supplementary Table 4**). Of note, these drugs/chemical compounds may have therapeutic value for depression or may increase depression risk due to their side effects, as the direction of their effects on target genes was not evaluated.

Taken together, our results revealed spatially patterned associations between glu-neurons and traits in the PFC lobe. We found that glu-neurons near the medial side of the PFC 14r (gyrus rectus) region were strongly associated with depression, with genes highly expressed in this area enriched in neural plasticity-related pathways and targets of psychiatric drugs.

## Discussion

In this study, we introduced gsMap, a method that integrates cellular gene expression profiles, cell spatial coordinates, SNP-to-gene linking maps, and GWAS summary data to spatially map cells to human complex traits. The gsMap method has been implemented in a Python package and is freely available at https://github.com/LeonSong1995/gsMap. Through extensive benchmark analyses with real ST and both real and simulated GWAS datasets, we demonstrated that gsMap was accurate and powerful in spatially aware discovery of associations between traits and cells. By applying gsMap to high-resolution brain ST datasets, we generated trait-brain maps that detail associations with 33 complex traits at spatially resolved single-cell resolution. These maps cover both evolutionarily conserved brain regions (e.g., hippocampus) and advanced brain regions (e.g., cerebral cortex), and various complex traits related to cognition, emotion, and behavior. We have developed an interactive web tool to visualize and download these trait-brain association maps, which are available at https://yanglab.westlake.edu.cn/gsmap/home.

Cells in the brain are usually assigned to conventional cell types (e.g., glu-neurons) to study their relevance to diseases^14,22,55,56^. However, our results suggest that due to within-cell type heterogeneity, only assigning cells to such cell types is not sufficient to understand their roles in diseases. We show that cells of the same conventional cell type in the brain exhibit a wealth of heterogeneity regarding their relevance to traits or diseases, and such heterogeneity is correlated with their spatial distributions. Our findings, in conjunction with previous morphology, electrophysiology and RNA sequencing studies^32,57-61^, indicate that spatially patterned within-cell type heterogeneity might be the basic organization rule for cells in the mammalian brain. Why do cells in the mammalian brain exhibit spatially patterned heterogeneity? Possible explanations include, first, that the spatial heterogeneity of brain cells is associated with the brain development process^62^. During brain development, cell differentiation and migration result in the establishment of distinct brain regions with specific cellular properties. This spatial patterning ensures that different brain areas are equipped to carry out specialized functions. Second, the spatial heterogeneity of brain cells is a natural reflection of the complex brain functions. The spatially patterned heterogeneity of brain cells increases diversity yet still ensures the organized harmony of cell elements in neural circuits, allowing for a more nuanced and sophisticated information processing in the mammalian brain^63,64^.

Several of our findings hold clinical value. First, we consistently observed distinct spatial patterns of glu-neurons related to cognitive and mood traits. Notably, SCZ-associated glu-neurons strongly overlapped with those associated with cognitive traits such as IQ. This finding suggests that the impairment in the brain responsible for cognition may be one of the major pathological changes in SCZ patients, elucidating why disorganized thinking is one of the most prevalent symptoms in SCZ ^65,66^. Second, our results highlighted several brain regions with potential therapeutic value. We demonstrated that the glu-neurons in the dorsal side of CA1 exhibited a strong association with SCZ, characterized by elevated expression of calcium signaling-related genes. Glu-neurons in the midbrain, CA1 superficial side, and gyrus rectus medial side were highly associated with depression, characterized by increased expression of neural plasticity-related genes. These results provide insights into the development of targeted therapeutics for complex psychiatric disorders. The strong enrichment of psychiatric drug targets in the gyrus rectus medial side further strengthens the clinical relevance of the identified brain regions.

We note several limitations in our study, which could serve as potential avenues for future research. First, owing to limitations in the presently available human ST data, we relied on brain ST data obtained from mice and macaques to spatially map brain cells associated with human complex traits. Despite the replication of our findings across diverse ST datasets and the demonstration of consistent gsMap results between the macaque PFC and the human DLPFC, the use of non-human ST data inevitably leads to a reduction in statistical power due to inherent differences between humans and these animal models, though the use of GWAS summary data from large-scale studies may partially offset this power loss. Given the rapid development of spatial omics technologies and the reduction in associated costs^4,5,67^, we anticipate the generation of extensive, high-quality spatial omics data using human tissues in future studies. These datasets would serve as valuable resources for further validating our results and making novel discoveries that cannot be achieved using non-human ST data. Second, associations between traits and cells identified by gsMap do not imply a causal relationship. Because of the correlation of gene expression profiles among cells, it is possible for cells that do not biologically contribute to a trait to be identified as associated with the trait. For example, we detected associations of root ganglion cells with SCZ using the mouse embryonic ST data, which might be caused by the correlated gene expression values between root ganglion cells and brain neurons. Third, our current gsMap method analyzes each ST section separately, potentially overlooking biological heterogeneity among sections, particularly in datasets containing serial tissue sections. Future work is warranted to enhance the method to handle multiple ST sections simultaneously, thereby capturing biological variances across these sections. Finally, the primary focus of this work was to apply gsMap to explore the spatial distribution of trait-associated cells in the brain, leaving the exploration of other tissues for further investigation.

## Methods

### Ethical approval

This study was approved by the Ethics Committee of Westlake University (approval no. 20200722YJ001).

### gsMap method

#### Design principles

To effectively illustrate gsMap, we first summarize its design principles. gsMap utilizes the framework of S-LDSC^19^ to assess whether genetic variants, mainly SNPs, located in or near genes specifically expressed in a spot in ST data are enriched for genetic associations with a trait of interest. To precisely estimate gene expression specificity for individual spots, gsMap aggregates information from homogeneous spots, a crucial step given the sparsity and high technical noise in gene expression profiles of individual spots in ST data^15,68,69^. Using spatial coordinates alone is inadequate to identify homogeneous spots because spatially neighboring spots may not necessarily belong to the same cell type. Using gene expression profiles alone could also lead to biased identification of homogeneous spots due to technical noise. To address these limitations, gsMap employs a GNN to learn embeddings that integrate spatial coordinates and gene expression profiles, and then identifies homogeneous spots for each focal spot based on their similarity in the embedding matrix. gsMap then estimates gene specificity scores for each focal spot by aggregating information from its homogeneous spots.

#### Data input

gsMap requires inputs of 1) GWAS summary statistics, 2) sequencing-based ST data comprising transcriptome-wide gene expression profiles and spatial coordinates of individual spots, 3) linkage disequilibrium (LD) reference data, and 4) optionally, SNP-to-gene linking maps. We noted that the resolution of ST data varies significantly across different platforms, ranging from one spot on sequencing array chips representing a cluster of cells (e.g., 10X Visium) to spots at sub-cellular resolution (e.g., Stereo-seq). To ensure the robustness of gsMap when handling ST data at sub-cellular resolution, cell segmentation^70^ analysis is required, which merges original spots on sequencing array chips into individual cells. For simplicity, we continue to use spots to denote data points in ST data, where one spot represents an individual cell (after cell segmentation analysis) in high-resolution ST platforms or multiple cells in conventional ST platforms.

#### Processing gene expression data

The gene expression count matrix is log-transformed and normalized according to the library size of each spot. Subsequently, the normalized gene expression matrix is standardized to attain a zero mean and unit variance. From this standardized matrix, the top *h* (default, 3000) highly variable genes (HVGs) are selected. The resulting processed gene expression matrix is represented as **X** ∈ ℝ^*n*×*h*^, with *n* denoting the number of spots and *h* representing the number of selected genes.

#### Building a spatial graph of the spots

The spatial coordinates of individual spots are transformed into an undirected graph, denoted as *G* = (*V, E*). In this graph, each vertex *v* ∈ *V* represents a spot, while *E* represents the set of connected edges between spots. For balancing the performance and computational efficiency, gsMap, by default, considers the 10 nearest neighboring spots for each spot, determined by their Euclidean distance in spatial coordinates. The resulting graph can be delineated by the adjacency matrix **A** ∈ ℝ^*n*×*n*^ with *n* denoting the number of spots. If spots *i* and *j* are connected in the graph, *A*_*ij*_ = 1; otherwise, *A*_*ij*_ = 0.

#### Learning embedding matrix

The standardized gene expression matrix **X** and the spot spatial graph ***A*** are then integrated into a embedding matrix **Z** ∈ ℝ^*n*×*m*^, where *m* represents the number of features (set as 32), using the graph attention (GAT) auto-encoder framework. The advantage of GAT^71^ lies in its trainable edge weights among connecting spots, enabling higher weights for spots with analogous gene expression patterns during information aggregation. A detailed description of the graph attention auto-encoder can be found in the **Supplementary Note 1**.

The loss function of the graph attention auto-encoder in gsMap includes two parts: 1) the mean squared error used for reconstructing gene expression matrix and 2) the cross-entropy loss employed for predicting cell types of spots:

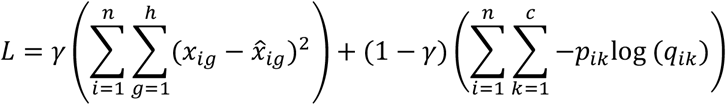

where *x*_*ig*_ represents the normalized expression value of spot *i* for gene *g* with 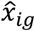 being the reconstructed value, *p*_*ik*_ represents the probability of spot *i* belonging to cell type *k* with *q*_*ik*_ being the predicted probability, *c* represents the number of cell types, and *γ* is the hyperparameter that balances the reconstruction loss and cross-entropy loss. In practice, *γ* is set to 0.5 in most cases. In scenarios where annotated cell types are unavailable, *γ* is set to 1. During the training process, the Adam optimizer^72^ is utilized to minimize the loss function and the exponential linear unit (ELU)^73^ is employed as the activation function. The weight decay is set to 1e-4, and the maximum number of iterations is set to 1000. The iteration is considered as converged when |*L*^*t*+1^ − *L*^*t*^| < 10^−4^.

#### Identification of homogeneous spots

The embedding matrix **Z** integrates information from gene expression values, spatial locations of spots, and cell type priors. We then identify homogeneous spots for each focal spot based on their cosine similarity in this latent space:

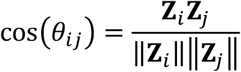

where **Z**_*i*_ ∈ ℝ^*m*×1^ is the embedding vector of spot *i*. For each focal spot, we select the top *d* spots that show the highest cosine similarity with it. This process aligns each individual spot with *d* spots that are spatially close and share similar transcriptomic profiles, referred to as the micro-domain (*D*). In this study, we set *d* to 20 for ST data generated from the 10X Visium platform and 50 for ST data generated from the Stereo-seq platform.

#### Estimation of gene specificity score

We rank genes in individual spots based on their expression levels, with higher expression values receiving higher ranks. We opt to use gene expression ranks instead of gene expression values because the former demonstrates greater robustness to technical noise^74^. For each gene, its expression specificity within each focal spot is assessed by calculating the geometric mean of its expression rank across the micro-domain of the focal spot, divided by the geometric mean of its expression rank across all spots in the ST data. For gene *g, F*_*ig*_ represents its expression specificity in spot *i*, calculated as:

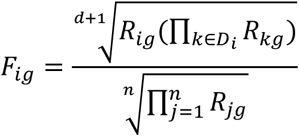

where *R*_*ig*_ denotes the expression rank of gene *g* in spot *i*, and *D*_*i*_ represents the micro-domain (i.e., set of homogonous spots) of spot *i* with *d* denoting the spot number. If *F*_*ig*_ < 0, indicating that this gene is not specifically expressed within the focal spot, we set it to 0. Additionally, we compare the expression proportion of each gene across the focal spot micro-domain to its proportion across all spots. If this ratio is smaller than 1, suggesting that the large *F*_*ig*_ might be due to outliers with discordant high expression ranks, we also set *F*_*ig*_ to 0. To align the scale between GWAS summary statistics and the estimated gene specificity, we project it onto the Gaussian kernel to further differentiate genes with high expression specificity:

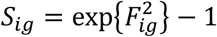

where *S*_*ig*_ denotes the final specificity score of gene *g* in spot *i*.

#### Mapping gene specificity score to SNPs

We employ the standard 100kb gene window-based approach to map the specificity score of each gene to the corresponding SNPs. Additionally, we provide an option to incorporate SNP-to-gene linking maps established from epigenomic data (e.g., Roadmap^17^ and Activity-by-Contact model^18^) for mapping SNPs located outside the 100kb window to genes. This process yields a unique set of SNP annotations for each spot.

#### Linking genomic annotations with GWAS data

Treating each spot as a set of SNP annotations, gsMap assesses whether SNPs with higher GSS are enriched for heritability for the trait of interest using the S-LDSC^19^ framework, conditional on the baseline SNP annotations. The S-LDSC in gsMap can be considered as a linear regression analysis between GWAS χ^2^ statistics and stratified LD scores computed using SNP annotations from individual spots. A detailed explanation of S-LDSC can be found in the **Supplementary Note 2**. The LD reference data used in this study were obtained from the 1000 Genomes Project Phase 3 (1KGP3)^75^. Following previous studies^14,76^, we use block-jackknife to estimate the standard error of the regression coefficient in S-LDSC. *P* value is computed using a one-sided Z-test, assessing whether the regression coefficient is significantly larger than 0. A smaller *P* value indicates a stronger relevance of the focal spot to the trait of interest.

#### Estimating the strength of enrichment for a spatial region

To evaluate relevance between a specific spatial region and the trait of interest, gsMap employs the Cauchy-combination test^21^ to aggregate *P* values of individual spots within the spatial region:

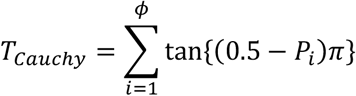

where *P*_*i*_ represents the *P* value of spot *i* belonging to the spatial region. The aggregated *P* value for the spatial region is approximated as:

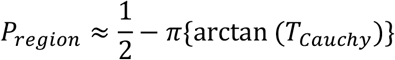

#### Running time

We have optimized the gsMap code to ensure its efficiency in handling large ST data. A summary of the gsMap runtime for each step is available in the **Supplementary Note 3**.

### Quantifying the association of a spatial region with a trait relative to all other regions

We employed an odds ratio (OR), computed as the ratio of trait-associated cells to non-associated ones in a specific region divided by the ratio in all other regions, to compare gsMap results of a specific spatial region across traits with varied GWAS statistical power. This metric quantifies the strength of a region’s association with a trait, relative to all other regions. Consequently, it compensates for differences in gsMap results due to variations in GWAS statistical power, allowing for a more meaningful comparison of gsMap results across traits. The significance of the OR was evaluated using a chi-squared test.

### Simulations

We simulated null scenarios where causal SNPs are randomly distributed across the genome, using real genotype data on 100K unrelated individuals of European ancestry from the UKB^24^. We used the HapMap3 SNPs and filtered out SNPs with a minor allele frequency (MAF)<0.01 or Hardy-Weinberg equilibrium (HWE) test *P* value<10e-6, resulting in a total of 1,195,548 SNPs. We employed GCTA (V1.94.1)^77^ to generate quantitative traits based on the real genotype data varying polygenicity (i.e., proportion of SNPs being causal) and heritability (i.e., proportion of variance in the phenotype attributed to the causal SNPs). Causal SNPs were varied from 10K to 500K, randomly sampled across the entire genome, and the heritability was varied from 0.1 to 0.6. Each simulation was replicated three times, with 36 simulation replicates in total. We used PLINK (V1.90)^78^ to associate SNPs with the simulated phenotypes with the first 10 principal components, derived from SNPs, fitted as covariates.

### GWAS summary statistics

We analyzed GWAS summary statistics for 110 complex traits, including diseases, from the UKB and other publicly available sources (average N=385K, **Supplementary Table 1**). All GWASs were well powered, as reflected by LDSC estimates of heritability with chi-squared statistics of at least 25. We excluded the major histocompatibility complex (MHC) region from all analyses due to its complexity^19^.

### Spatial transcriptomics datasets

We included four spatial transcriptomics datasets in this study: the mouse embryonic dataset^6^ (generated by Stereo-seq), mouse brain dataset (generated by Stereo-seq)^6^, macaque cortex dataset (generated by Stereo-seq)^8^, and human DLPFC dataset (generated by 10X Visium)^9^. To align mouse or macaque genes with human genes, we utilized the biomaRt (V3.18)^78^ R package to conduct homologous gene transformations. The average gene numbers after homologous transformation are 16,330 for the mouse datasets and 13,536 for the macaque dataset. Following the standard analysis pipeline, we utilized the Scanpy (V1.9.6)^79^ Python package to process each ST dataset. Details of each dataset are summarized below.

#### Mouse embryonic dataset

We analyzed 54 coronal sections from the mouse embryonic dataset with an average of 81,125 spots per section, spanning from the embryonic stage of E9.5 to E16.5^6^. Data at both bin50 resolution (53 sections) and single-cell resolution (1 section) were included in this study. We obtained access to the h5ad files, each including the gene expression count matrix, cell type annotations, and spots spatial coordinates for a section. We validated the cell type annotations based on known marker genes.

#### Mouse brain dataset

We analyzed two sections from the mouse brain dataset: one coronal section from adult mouse brain (50,140 spots) and one sagittal section from E16.5 embryonic mouse brain (65,303 spots)^6^. Both sections are at single-cell resolution. We had access to the h5ad files and verified the cell type annotations using known marker genes.

#### Macaque cortex dataset

We analyzed 162 coronal sections from the adult macaque cerebral cortex dataset with an average of 266,654 spots per section^8^. All sections are at single-cell resolution. We obtained access to the SCT-transformed^80^ gene expression matrices and the metadata files. For each section, we aligned the spots spatial coordinates, cell type annotations, cortex region annotations, and section-cutting positions (EBZ) and compiled all the aforementioned information into an h5ad file.

#### Human DLPFC data set

We analyzed 8 sections from the adult human DLPFC dataset with an average of 3,973 spots per section^9^. These data were generated using 10X Visium, with each spot containing a few dozen cells. We had access to the h5ad files, each including the gene expression count matrix, spots spatial coordinates, and cortex layer annotations.

### Comparison of the gsMap results from the human and macaque datasets

We applied gsMap to 8 human DPLFC ST sections and observed highly consistent results across these sections. We then calculated an OR for each cortex layer, representing the strength of association of a cortex layer with a trait, relative to all other layers. To ensure a robust comparison between the human and macaque results, we compared the median OR value of each cortex layer across 8 human DPLFC ST sections to that across 9 macaque PFC ST sections. Considering that there were only 5 matched cortex layers between the human and macaque datasets, we pooled the OR values from 22 brain-related traits, resulting in 110 data points in the comparison analysis.

### Integration of GWAS summary statistics with ST data using scDRS

scDRS^13^ is a method that can integrate GWAS summary statistics with scRNA-seq data to identify cells relevant to a trait. Briefly, scDRS computes a trait-enrichment score to examine whether a cell has excess expression levels across a set of trait-associated genes. These genes were derived from GWAS summary statistics using gene-based association tests (e.g., MAGMA^81^). To assess the statistical significance of the trait-enrichment score, a *P* value is calculated by comparing the trait-enrichment score to those computed from control genes with matched expression characteristics. Though originally developed for scRNA-seq data, scDRS can, in principle, be applied to ST data by regarding each spot in the ST data as a cell. Following the standard analysis protocol of scDRS, we first used MAGMA to generate gene-based test statistics from the GWAS summary statistics, utilizing reference LD data obtained from 1KGP3, as done in gsMap analysis. Next, we used the ‘munge-gs’ command in scDRS and set the ‘n-max’ parameter to 1000, to generate a gene-weight file for the top 1000 trait-associated genes identified from the MAGMA analysis. Finally, we used the ‘compute-score’ command in scDRS and set the ‘n-ctrl’ parameter to 1000, to obtain the trait-enrichment *P* value for each spot.

### Psychiatric drug targets

We collected drug target genes from the DrugBank database^52^ and Drug Repurposing Hub database^51^. For the DrugBank database, we selected drugs intended to treat psychiatric disorders categorized under the International Classification of Diseases (ICD) codes F00 to F99. From the Drug Repurposing Hub database, we selected drugs categorized under the psychiatric disease areas. In total, we identified 417 target genes from drugs that are either approved or undergoing clinical trials for treating psychiatric disorders. Based on the identified drug target genes, we used Fisher’s exact test to assess whether genes highly expressed in the macaque PFC 14r region are enriched in these drug target genes, compared to all detected genes, or genes highly expressed in other PFC regions. The module score of these 417 drug targets genes for individual spots was computed using the ‘AddModuleScore’ function in the Seurat (V4.4.0) R package with the default settings^53,54^

### Gene ontology enrichment

We performed gene ontology (GO) enrichment analysis using the clusterProfiler^82^ (V3.18) R package with the default settings.

### Genetic correlation

We used the bivariate LDSC^76,83^ (V1.01) to estimate genetic correlations between trait pairs. The reference LD data used in the genetic correlation analysis was generated from 1KGP3.

### Statistics and reproducibility

We analyzed only existing datasets without employing any statistical methods to predefine the sample size. There was no exclusion of data from our analyses. Our study did not involve a design requiring randomization or blinding. We validated our results by performing the same analyses on independent datasets, and all replication analyses were successful.

## Data availability

This study used GWAS summary statistics for 110 traits, as summarized in **Supplementary Table 1**. The mouse embryonic and brain ST data are available at https://db.cngb.org/search/project/CNP0001543/. The macaque cortex ST data are available at https://db.cngb.org/search/project/CNP0002035/. The ST data for human DLPFC are available at https://research.libd.org/globus/. The reference LD data, generated from 1KGP3, are available at ftp://ftp.1000genomes.ebi.ac.uk/vol1/ftp/release/2013050. The baseline annotations of LDSC are available at https://data.broadinstitute.org/alkesgroup/LDSCORE. The DrugBank database is available at https://go.drugbank.com/. The Drug Repurposing Hub database is available at https://www.broadinstitute.org/drug-repurposing-hub. The human, mouse, and macaque reference genome data are available at https://nov2020.archive.ensembl.org/index.html. The gsMap results for different traits and ST datasets can be visualized and downloaded at https://yanglab.westlake.edu.cn/gsmap/home.

## Code availability

The source code for gsMap is available at https://github.com/LeonSong1995/gsMap.

## Acknowledgements

We thank Bo Li and Qiufu Ma for their insightful suggestions in interpreting the results from the brain-related analyses, and Feifei Cheng, Shuli Liu, Guiying Dong, Shufeng Sun, and Yazhou Guo for their valuable discussions and assistance in writing the manuscript. We thank the Westlake University High-Performance Computing Center for their assistance in computing. This research was supported by the Leading Innovative and Entrepreneur Team Introduction Program of Zhejiang (2021R01013), “Pioneer” and “Leading Goose” R&D Program of Zhejiang (2022SDXHDX0001 and 2024SSYS0032), the National Natural Science Foundation of China (U23A20165), and the Westlake University Research Center for industries of the Future (WU2022C002 and WU2023C010).

## Author contributions

JY and LS conceived the study and designed the experiment. LS developed the methods with input from WC and JY. LS and WC developed the software tool. LS and WC curated the data and conducted all analyses under the guidance of JY. JH and MG developed the interactive web. JY supervised the project. LS and JY wrote the manuscript with the participation of WC. All authors reviewed and approved the final manuscript.

