## Supplementary Notes and Figures for "Spatially resolved mapping of cells associated with human complex traits"

Song *et al*.

**Contents**

**Supplementary Notes 1-3**

**Supplementary Figures 1-19**

**Supplementary Tables 1-4**

**Supplementary References**

**Supplementary Note**

### 1. Graph attention auto-encoder

***Encoder*.** The gene expression matrix $\mathbf{X} \in \mathbb{R}^{n\times h}$, where $n$ denotes the number of spots and $h$ denotes the number of genes, first undergoes $k$-th ($k \in \{1,2,..,L\}$) fully-connected linear layers.

$$\mathbf{B}^{(k)} = \sigma(\mathbf{B}^{(k-1)}\mathbf{W}_k)$$

where $\mathbf{B}^{(0)} = \mathbf{X}$ is the input gene expression matrix for the linear layer, $\mathbf{W}_k$ is the trainable weight matrix of layer $k$, and $\sigma$ is the ELU activation function. This process result in a condensed gene expression feature matrix $\mathbf{B}^{(L)} \in \mathbb{R}^{n\times f}$, with $f$ being the number of features.

Next, we leverage the graph attention (GAT) layer to aggregate information from spatially connected spots. The advantage of GAT layer lies in its trainable edge attention among connecting spots, enabling higher attention for spots with analogous gene expression patterns during information aggregation[1]. The $\mathbf{B}^{(L)}$ matrix undergoes $h$-th ($h \in \{1,2,..,S\}$) GAT layers to generate the final embedding. For spot $i$, its embedding vector is calculated as follows:

$$\mathbf{Z}_i^{(h)} = \sigma(\sum_{j\in\Omega_i} a_{ij}^{h}\,(\mathbf{Z}_i'^{(h-1)}\mathbf{Q}_h)$$

where $\mathbf{Z}_i^{(0)} = \mathbf{B}_i^{(L)} \in \mathbb{R}^{f\times 1}$ is the input matrix for the GAT layer, derived from the previous linear layer, $\mathbf{Q}_h$ denotes the trainable weight vector at the GAT layer $h$, $a_{ij}^{h}$ is the edge attention value between spots $i$ and $j$ at layer $h$, $\Omega_i$ represents all spots connected with spot $i$ in the graph, and $\sigma$ is the ELU activation function. The edge attention value between spots $i$ and $j$ is computed as follows:

$$\mathrm{e}_{ij}^{(h)} = \sigma(\left[\mathbf{Z}_i'^{(h-1)}\mathbf{Q}_h \parallel \mathbf{Z}_i'^{(h-1)}\mathbf{Q}_h\right]\mathbf{V}^{(h)})$$

where $\mathbf{V}^{(h)}$ is the trainable weight vector and $\sigma$ is the LeakyReLU activation function. To ensure comparability edge attentions among spots, we normalized them as follows:

$$a_{ij}^{(h)} = \frac{\exp\,(\mathrm{e}_{ij}^{(h)})}{\sum_{j\in\Omega_i}\exp\,(\mathrm{e}_{ij}^{(h)})}$$

where $\Omega_i$ represents all spots connected with spot $i$ in the graph. In summary, the encoder uses linear and GAT layers to integrate the gene expression matrix and the spot spatial graph into the embedding matrix $\mathbf{Z}^{(S)} \in \mathbb{R}^{n\times m}$, where $m$ denotes the number of final features (set to 32 by default).

***Decoder*.** The decoder mirrors the configuration of the encoder, aiming to reconstruct gene expression matrix and predict cell types of spots, as described in the main text.

### 2. S-LDSC

After obtaining the SNP annotations by assigning GSS of each spot in ST data to SNPs, we employ the S-LDSC[2,3] framework to quantify the heritability enrichment of each spot for a complex trait using the following equation:

$$E(\chi_j^2) = N\left(l(j,s)\beta_s + \sum_c l(j,c)\beta_c\right) + 1$$

where $\chi_j^2$ denotes the association $\chi^2$ statistic of SNP $j$ with the trait; $N$ denotes the GWAS sample size; $l(j,s) = \sum_k a_{sk} r_{jk}^2$ is the stratified LD score of SNP $j$ with respect to SNP annotation $a_{s.}$ (i.e., GSS of spot $s$) with $\beta_s$ being the coefficient and $r_{jk}^2$ being the squared LD correlation between SNPs $j$ and $k$; $l(j,c) = \sum_k a_{ck} r_{jk}^2$ is the stratified LD score of SNP $j$ with respect to a baseline annotation $a_{c.}$ with $\beta_c$ being the coefficient. The baseline annotations include: 1) a constant value of 1 for all SNPs; 2) binary values (0 or 1) to indicate whether SNPs are mapped to genes; 3) optionally, binary values to indicate whether SNPs are mapped to other functional annotations, including coding, conserved, and regulatory regions (e.g., promoter, enhancer, histone marks). Following prior work[4], we use the block-jackknife to estimate the standard error of $\hat{\beta}_s$. Then, the $P$ value of $\hat{\beta}_s$ is computed using a one-sided Z-test, assessing whether it is greater than 0.

### 3. gsMap runtime

Below is the approximate runtime of each step in gsMap, using mouse E16.5 embryonic ST data with 120K spots as an example:

CPU: Intel Xeon Silver 4210R; GPU: NVIDIA V100.

1) Processing ST data and identifying homogeneous spots: 0.5 GPU hour or 2 CPU hours;
2) Generating gene specificity scores: 1 CPU hour;
3) Generating stratified LD scores: 1 CPU hour (for chromosome 1);
4) Linking stratified LD scores to GWAS data: 2 CPU hours.
5) Aggerating $P$ values using the Cauchy-combination test: 0.1 CPU hour.

Note that the outputs from steps 1 to 3 are adaptable for linking different GWAS summary statistics to the same ST data.

### Supplementary Figures

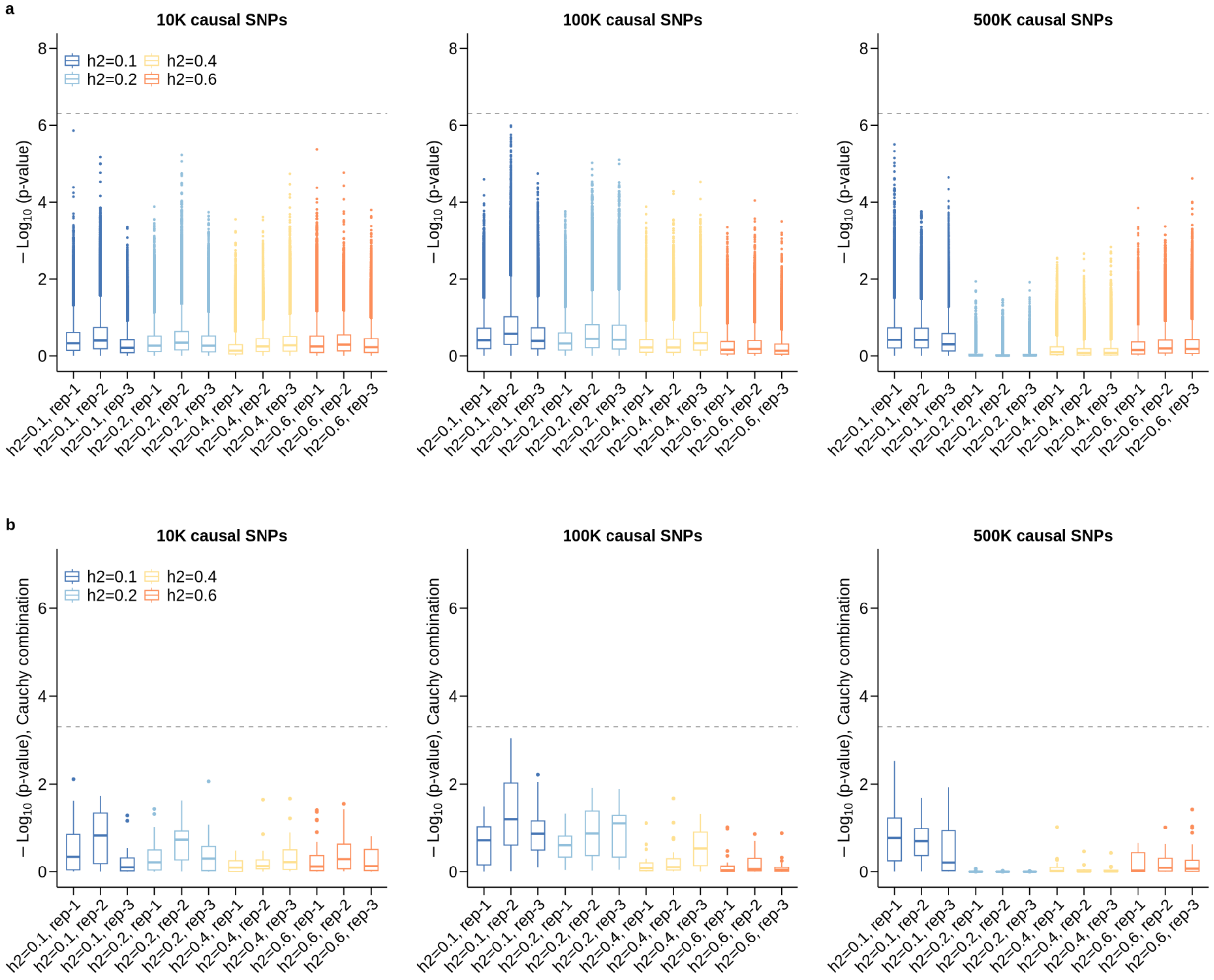


**Supplementary Figure 1. gsMap results in null simulations.** To validate that the false-positive rate in gsMap is well-controlled, we simulated null scenarios wherein the causal variants are randomly distributed across the genome, meaning that causal variants are not enriched in any gene regions. Simulations were performed using real genotype data on 100K unrelated individuals of European ancestry from the UKB. We simulated different scenarios with varied heritability and polygenicity. (a-b) Boxplot of gsMap results at the spot level (a) and region level (b) in null simulations with the number of causal SNPs set to 10K, 100K, and 500K. The x-axis displays trait heritability, varying from 0.1 to 0.6; the y-axis shows the association -log10(*P* value). In each box, the central line denotes the median, notches represent the 95% confidence interval (CI), the box indicates the interquartile range (IQR), and whiskers extend up to 1.5 times the IQR, with outliers shown as individual dots. The dashed line represents the threshold at FDR=0.05.

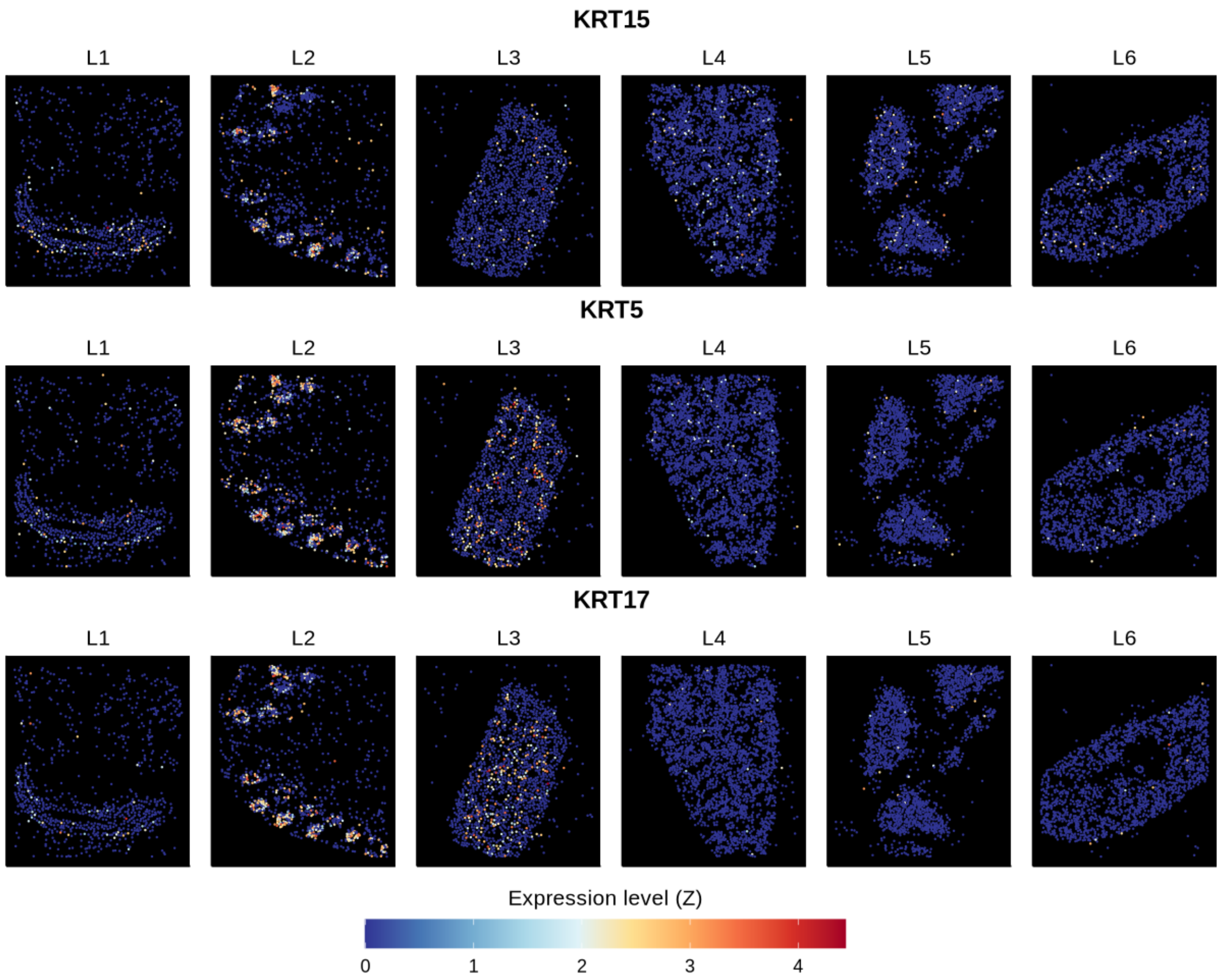


**Supplementary Figure 2. Expression pattern of hair follicle-associated marker genes**. The normalized expression levels of hair follicle-associated marker genes, including *KRT15* (Top), *KRT5* (Middle), and *KRT17* (Bottom), in six different spatial regions. Each point represents an individual epithelial cell, colored according to the normalized expression levels of the marker genes. The L2 region corresponds to the face region.

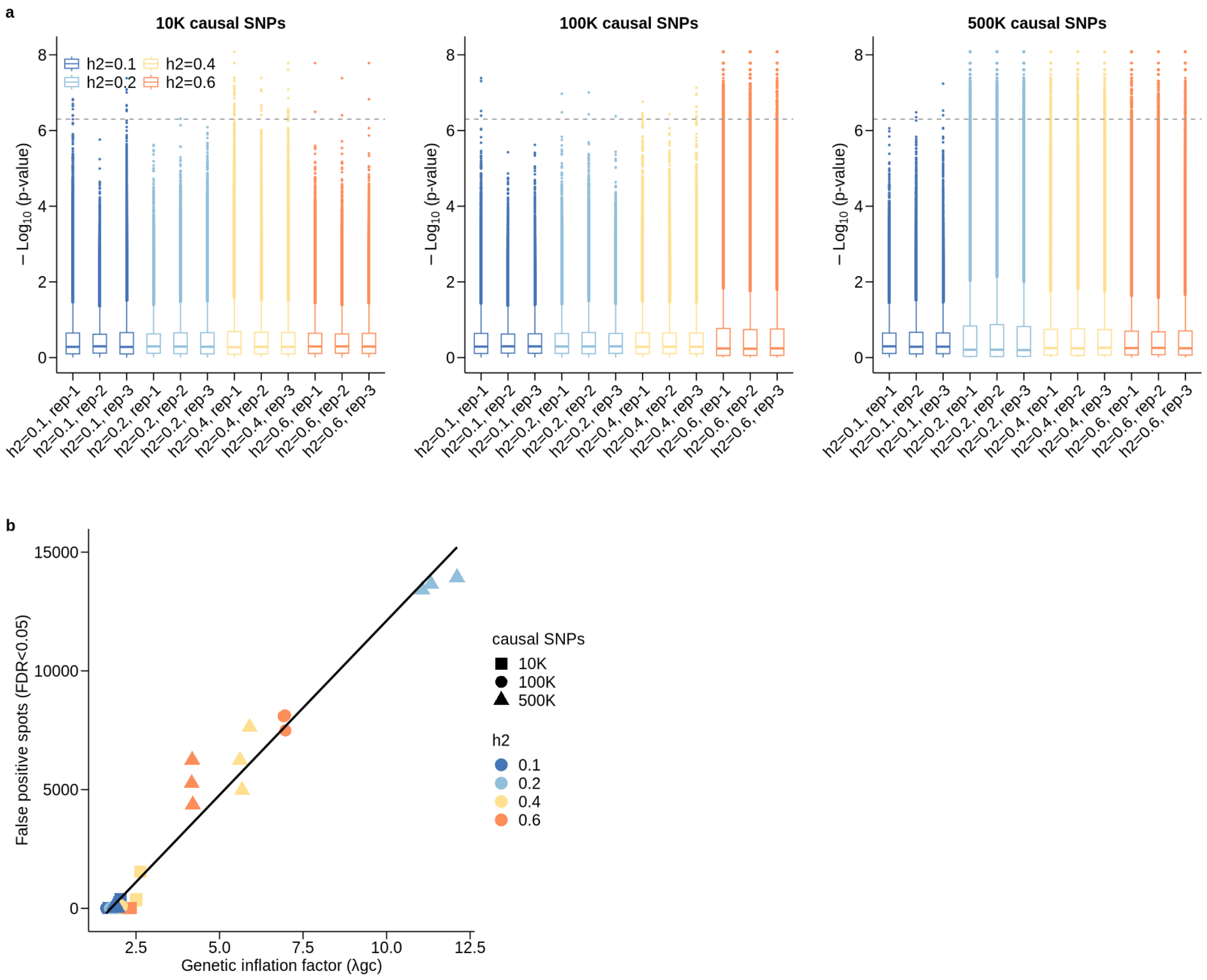


**Supplementary Figure 3. scDRS results in null simulations.** (a) Boxplot of scDRS results in null simulations with the number of causal SNPs set to 10K, 100K, and 500K. The x-axis shows trait heritability, varying from 0.1 to 0.6; the y-axis shows the association -log10(*P* value). In each box, the central line denotes the median, notches represent the 95% CI, the box indicates the IQR, and whiskers extend up to 1.5 times the IQR, with outliers shown as individual dots. The dashed line represents the threshold at FDR=0.05. (b) Scatter plot of the association between the number of false positive spots at FDR<0.05 from scDRS (y-axis) and the GWAS statistical power (x-axis). The genetic inflation factor, $\lambda_{GC}$, serves as an indication of the statistical power of the GWAS data. Each point represents one simulation replicate, with color and shape indicating the trait heritability and the number of causal variants, respectively.

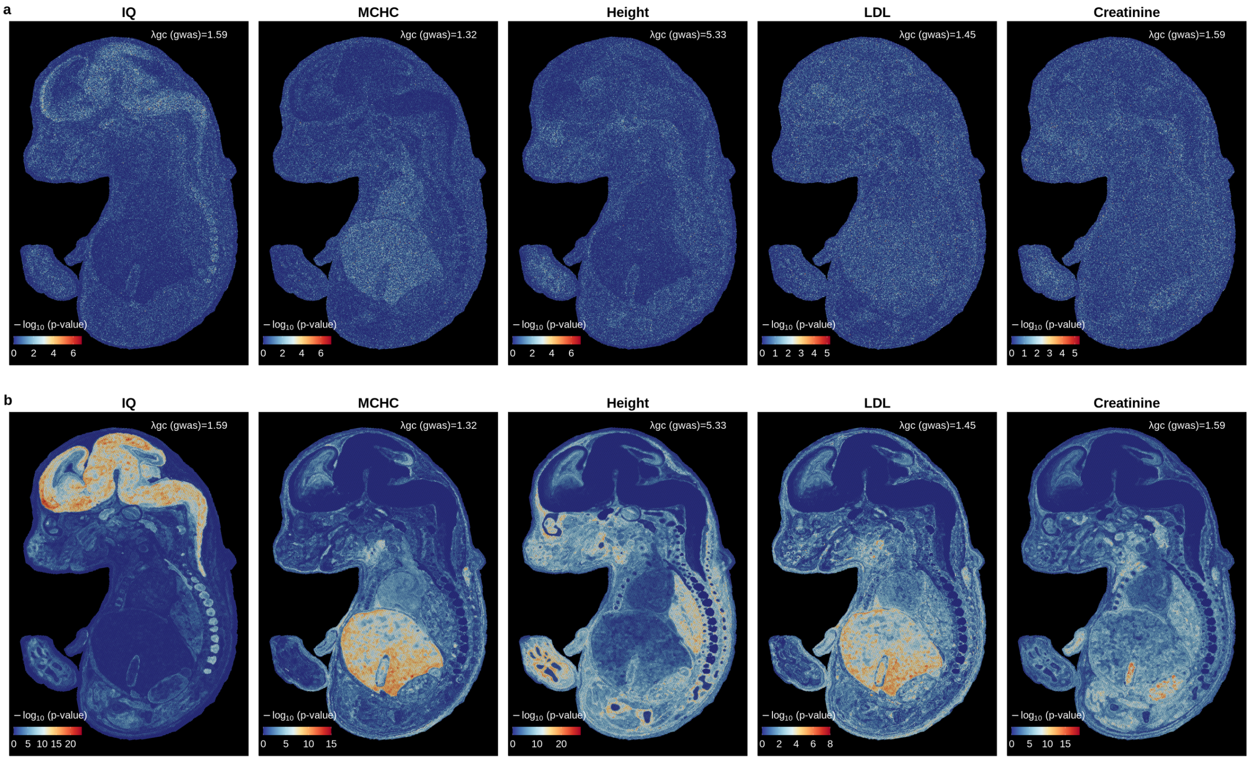


**Supplementary Figure 4. Comparison of scDRS and gsMap in real GWAS data analyses.** scDRS (a) and gsMap (b) results for IQ, MCHC, height, LDL, and creatinine using the mouse E16.5 embryonic ST data. Each point represents a spot, with the color indicating the significance of its association with a trait. The $\lambda_{GC}$ represents the genetic inflation factor of the GWAS summary statistics, serving as an indication of the power of the GWAS data.

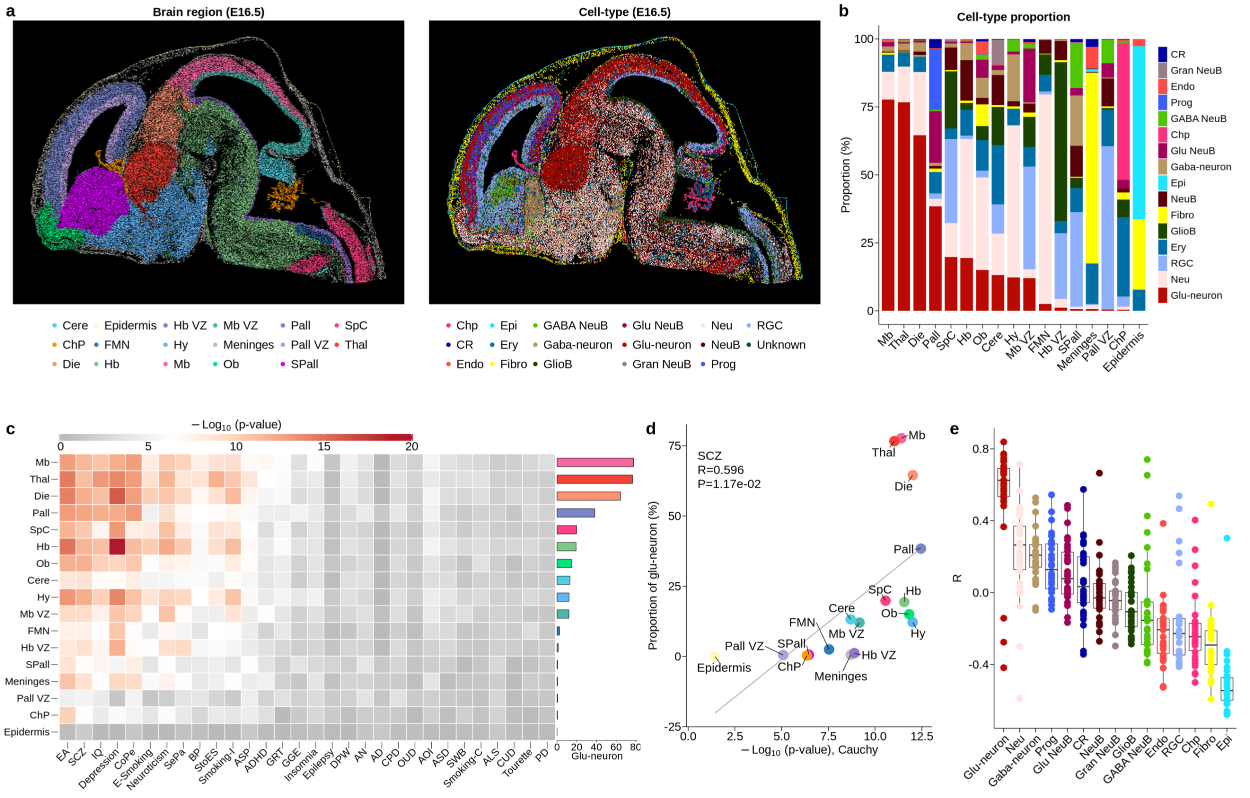

**Supplementary Figure 5. Genetically informed mapping of complex traits to mouse embryonic brain ST data.** (a) Mouse E16.5 embryonic brain ST data, where spots are colored by their brain regions (Left) or cell types (Right). Cere, cerebellum; Hb, hindbrain; VZ, ventricular zone; Pall, pallium; SpC, spinal cord; ChP, choroid plexus; FMN, facial motor nucleus; Hy, hypothalamus; Thal, thalamus; Die, diencephalon; Hb, hindbrain; Mb, dorsal midbrain; OB, olfactory bulb; SPall, subpallium. Neu, neuron; NeuB, neuroblast; Glu, glutamatergic; GABA, GABAergic; Fibro, fibroblast; Ery, erythrocyte; GlioB, glioblasts; CR, Cajal-Retzius cell; Epi, epithelial cell; Gran, granule; Prog, progenitor; Endo, endothelial cell; RGC, radial glia cell; Glu-neuron, glutamatergic neuron; Gaba-neuron, GABAergic neuron. (b) Bar plot of cell type proportions in each brain region. (c) Heatmap of associations of brain regions with different traits, with colors indicating the significance of the associations and the colored bar on the right representing the proportion of glu-neurons in each brain region. Each row corresponds to a specific brain region, and each column represents a trait. (d) Correlation between the significance of a brain region's association with SCZ (x-axis) and its proportion of glu-neurons (y-axis). The grey line represents the regression line. (e) Boxplot showing the correlation between significance of association with a trait and proportion of a cell type across brain regions. The x-axis displays different cell types; the y-axis shows the correlation coefficient. Each point represents a trait, with its color indicating the corresponding cell type.

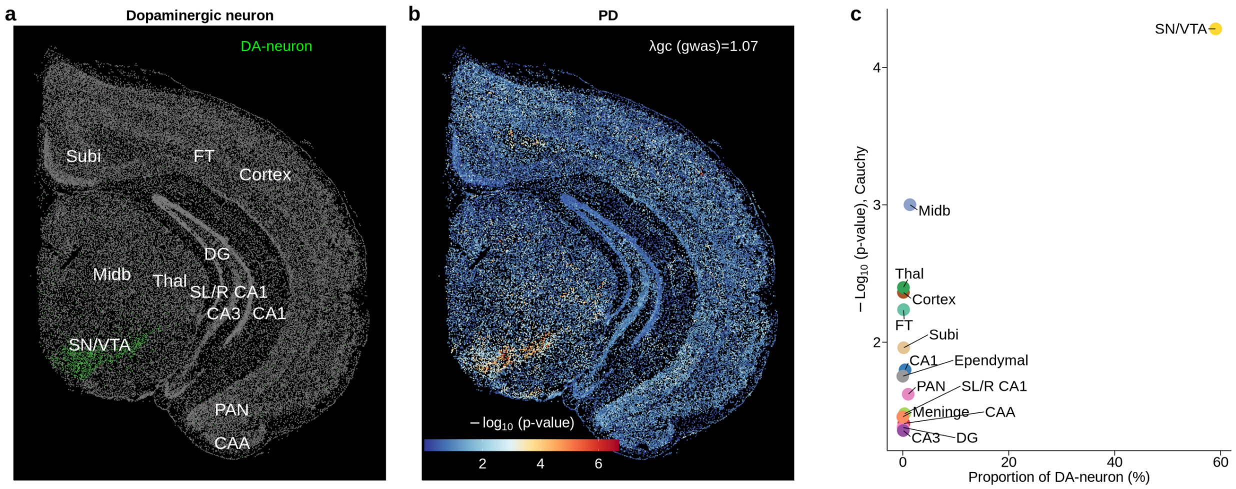


**Supplementary Figure 6. Genetically informed mapping of Parkinson's Disease to adult mouse brain.** (a) The distribution of dopaminergic neurons (DA-neurons) on the adult mouse brain ST data, where green points denote DA-neurons. CA1, cornu ammonis area 1; CA3, cornu ammonis area 3; CAA, cortical amygdala area; DG, dentate gyrus; FT, fiber tract; Midb, midbrain; PAN, posterior amygdala nucleus; SL/R CA1, stratum lacunosum/raditum cornu ammonis area 1; SN/VTA, substantia nigra/ventral tegmental area; Subi, subiculum; Thal, thalamus. (b) gsMap results for Parkinson's Disease (PD), where each point represents an individual cell colored by the significance of its association with PD. (c) Scatter plot showing proportions of DA-neurons in different brain regions (x-axis) and the significance of their associations with PD (y-axis).

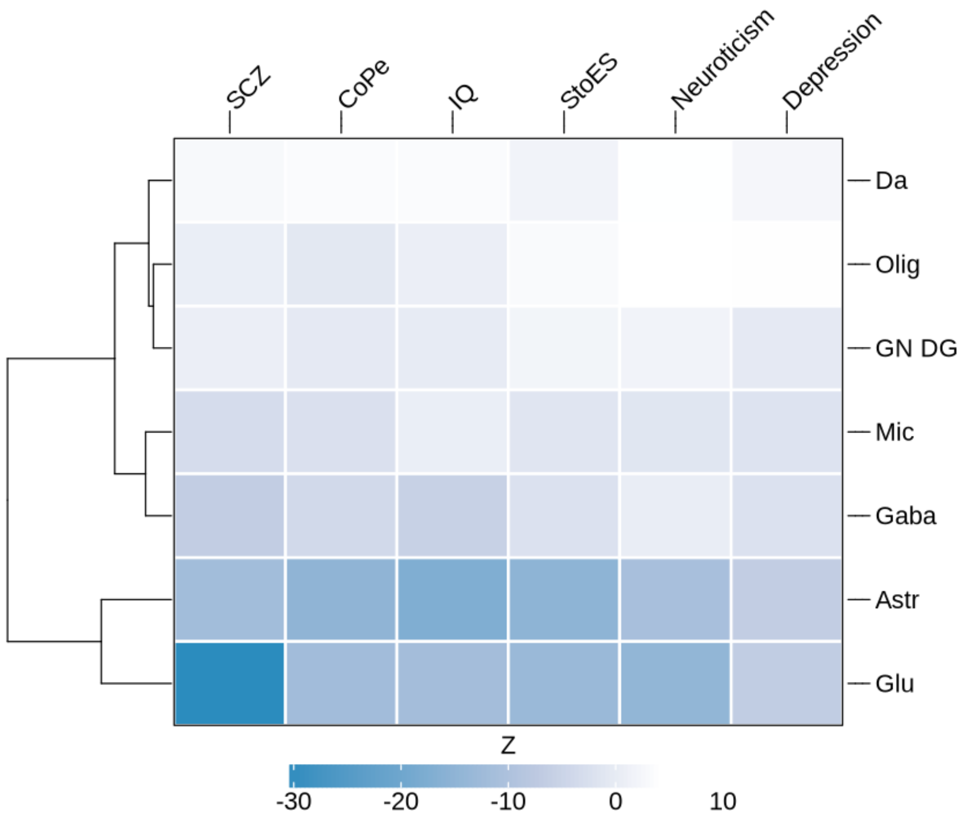


**Supplementary Figure 7. Correlation between the significance of associations of cells with a trait and their positions along the hippocampus CA1 D-V axis.** Heatmap of Z-scores for correlation between the significance of associations of a type of cells with a trait and their spatial positions along the hippocampus CA1 D-V axis. Each row corresponds to a cell type, and each column represents a trait.

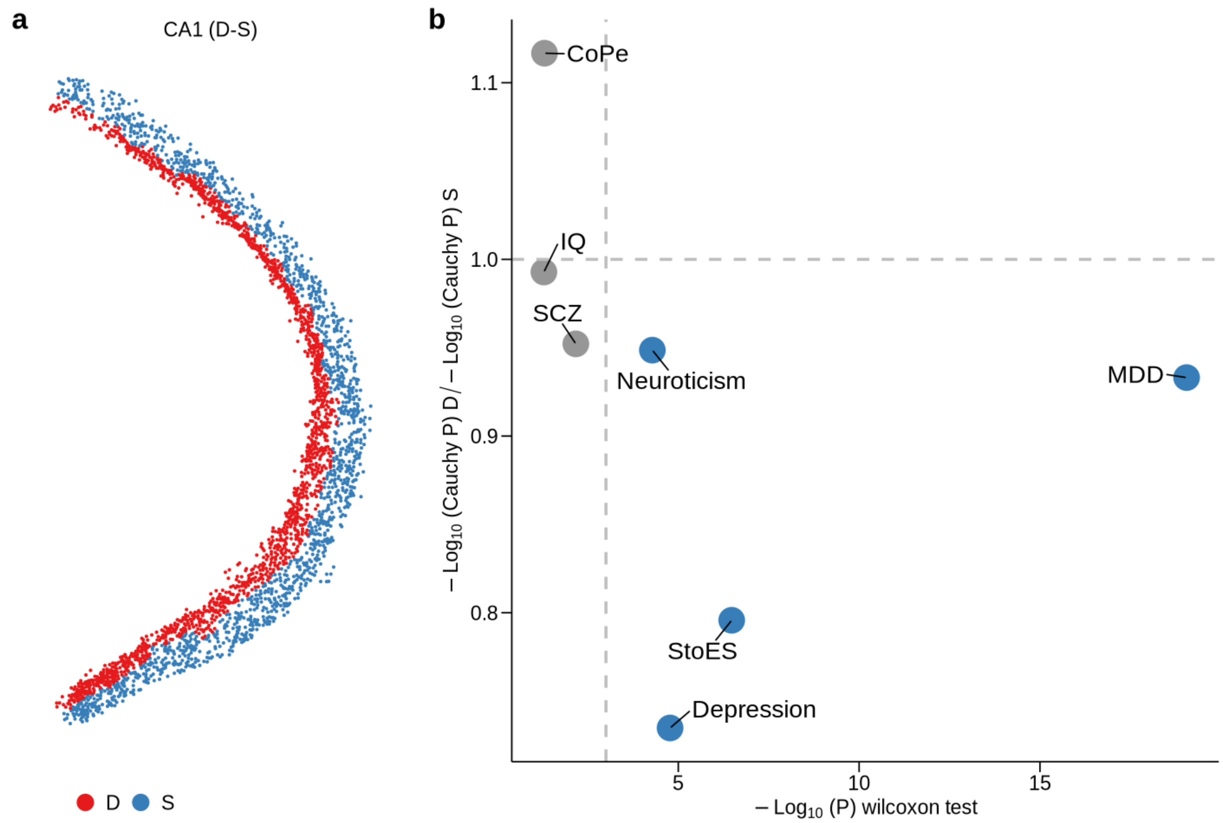


**Supplementary Figure 8. Spatial heterogeneity of glu-neurons along the hippocampus CA1 D-S axis.** (a) The hippocampus CA1 region was divided into the CA1 deep (D) side and the CA1 superficial (S) side along the D-S axis. (b) Comparison of gsMap results between the CA1 D side and the CA1 S side. The x-axis displays the Wilcoxon rank sum test *P* value to compare the significance of trait associations among glu-neurons in the CA1 D side and the CA1 S side. The y-axis displays the ratio of significance of association of glu-neurons in the CA1 D side with a trait, obtained using the Cauchy combination test, to that in the CA1 S side. Each data point represents a trait. The horizontal dashed line represents the ratio of 1, and the vertical dashed line indicates the significant threshold at FDR=0.01.

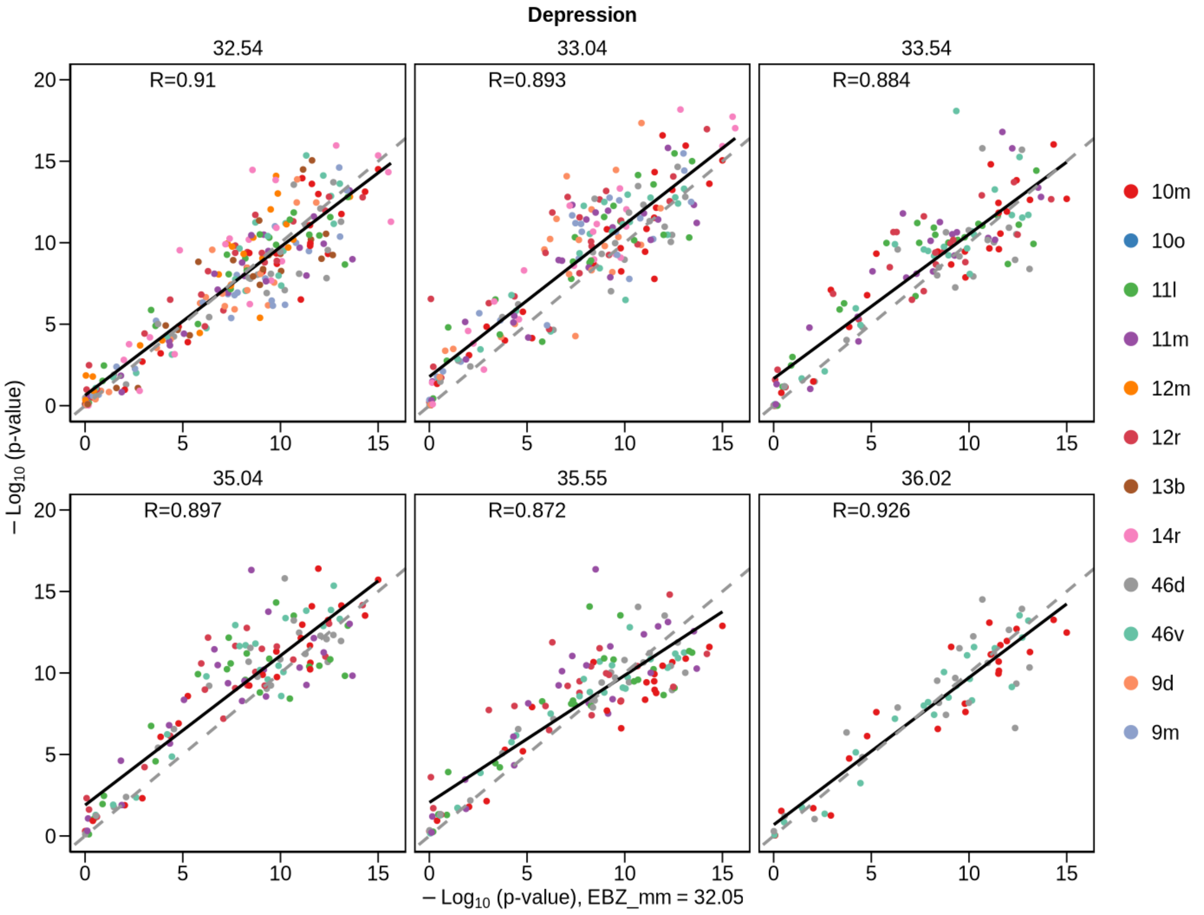


**Supplementary Figure 9. Associations of cell types in each of the cortex region with depression estimated using the EBZ 32.05 ST section vs. those estimated using the other ST sections.** The x-axis displays the associations of cell types with depression using the ST section of EBZ 32.05; the y-axis shows the corresponding associations estimated using the ST sections from EBZ 32.54 to EBZ 36.02. Each data point represents a cell type, with its color indicating the corresponding cortex region. The black line is the regression line, and the grey dashed line represents the line of equality.

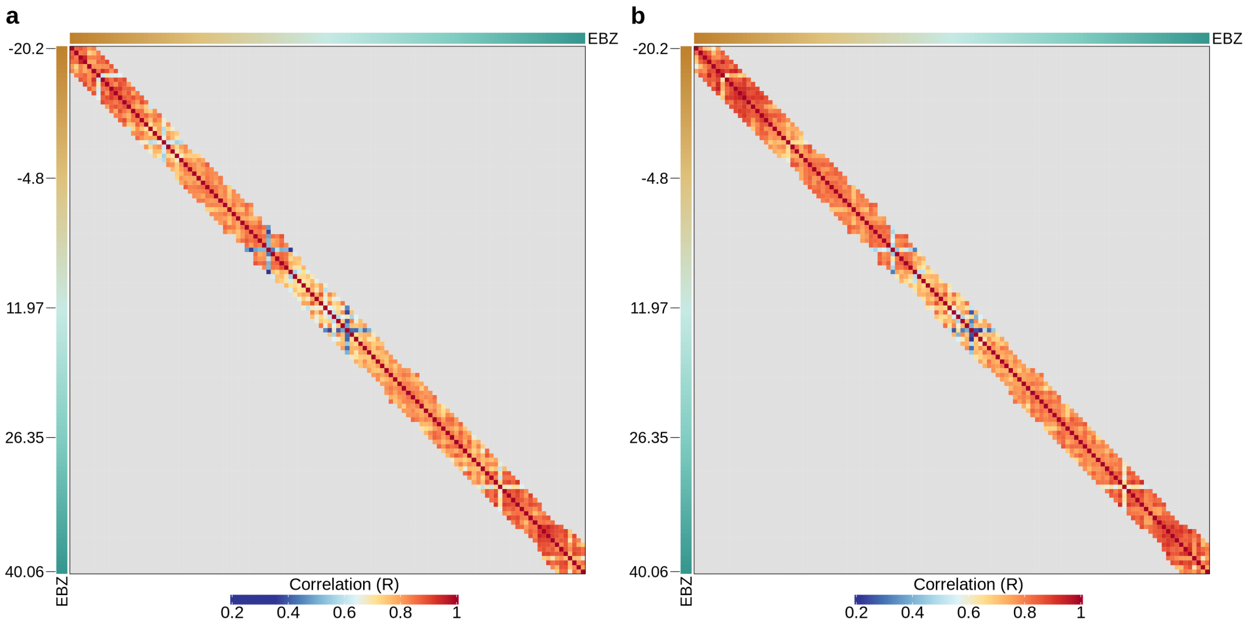


**Supplementary Figure 10. Correlation of gsMap results among different macaque ST sections.** Heatmap of Pearson correlations among gsMap results across 118 macaque cortex ST sections, spanning from the anterior cortex (EBZ 40.06) to the posterior cortex (EBZ -20.2), for SCZ (a) and depression (b). Each ST section was compared to its 10 adjacent ST sections. The row and column annotations of the heatmap indicate the cutting position of each ST section (EBZ, mm).

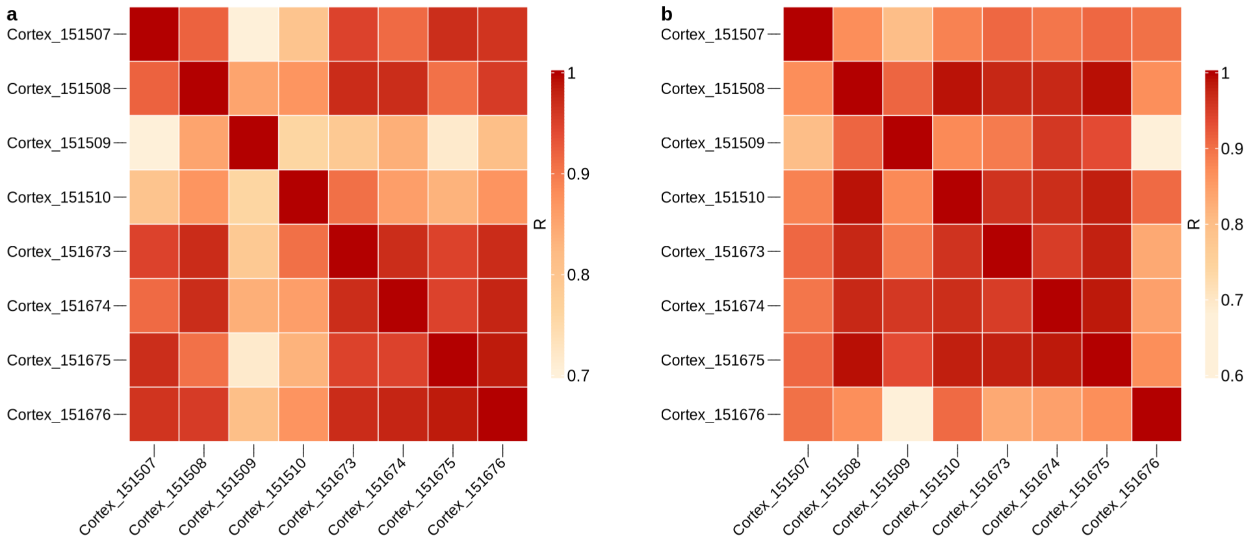


**Supplementary Figure 11. Correlation of gsMap results among different human DLPFC ST sections.** Heatmap of the Pearson correlations among gsMap results across 8 human DLPFC ST sections for SCZ (a) and depression (b).

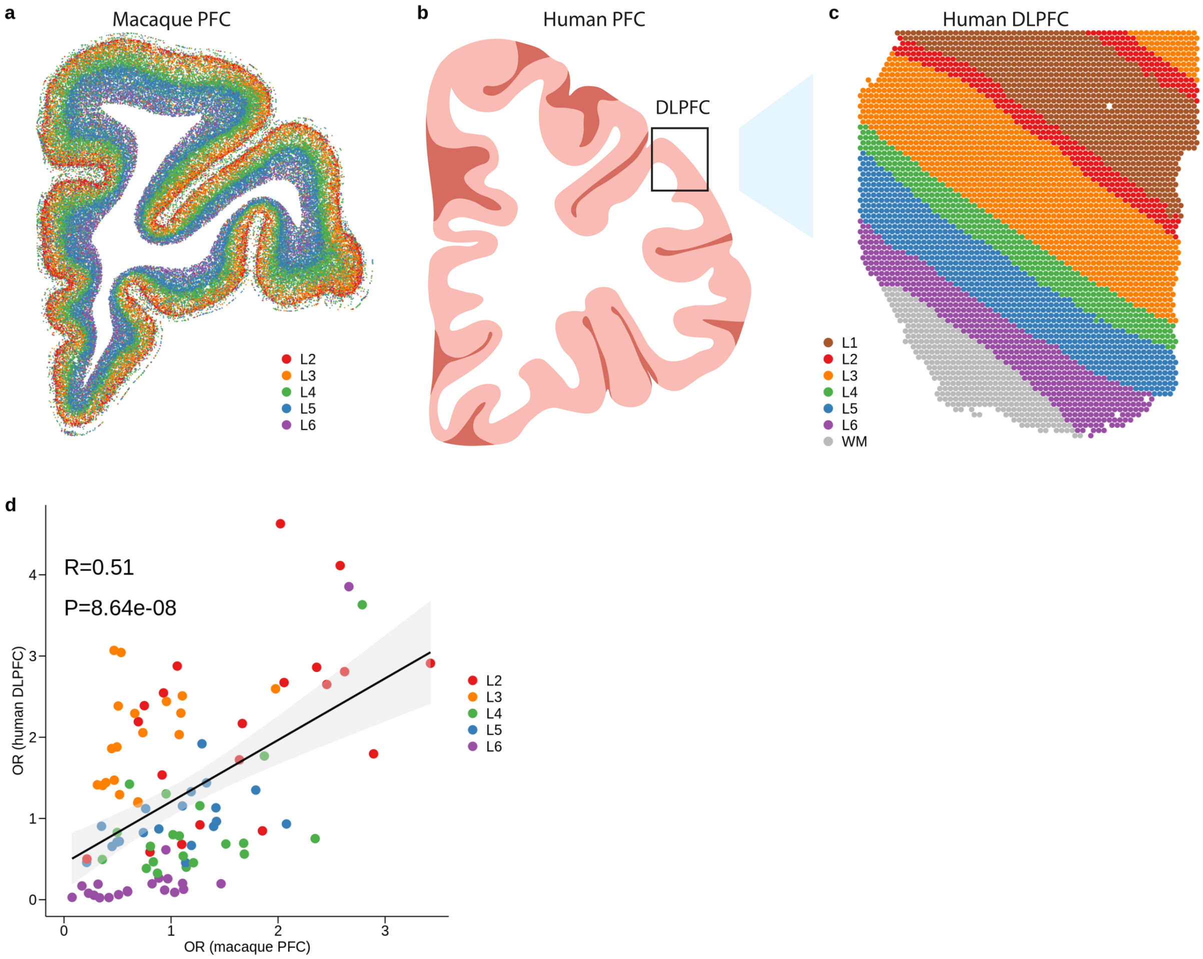


**Supplementary Figure 12. Comparison of gsMap results between human and macaque.** (a) Macaque prefrontal cortex (PFC) ST data. Each point represents an individual cell, with its color indicating the corresponding cortex layer. (b) Schematic of the PFC and DLPFC). (c) Human DLPFC ST data generated by the 10X Visium platform. Each point represents an individual spot (a few dozen cells), colored by its cortex layer. (d) Correlation between ORs of cortex layers estimated from the macaque PFC ST data and those estimated from the human DLPFC ST data. To ensure a robust comparison between human and macaque results, we compared the median OR of each cortex layer across 8 human DLPFC ST sections (y-axis) to that across 9 macaque PFC ST sections (x-axis). Considering that there were only 5 matched cortex layers between the human and macaque datasets, we pooled ORs from 22 brain-related traits. Each data point represents the OR of a cortex layer for a trait. The black line is the regression line, with the shaded area indicating the 95% CI.

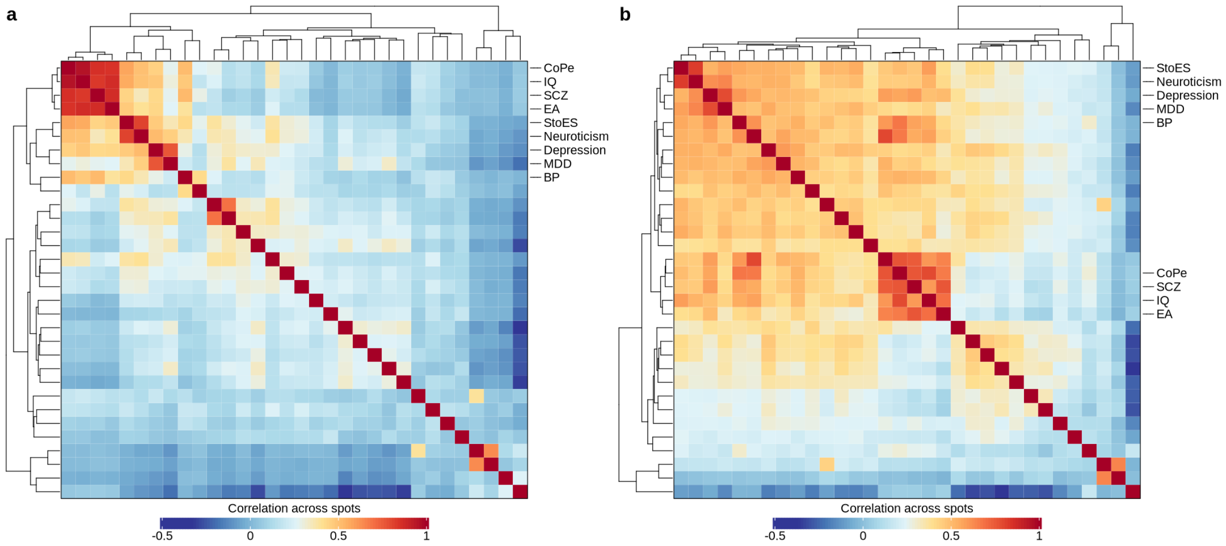


**Supplementary Figure 13. CRCs of brain-related traits estimated using mouse brain ST data.** Heatmap of CRCs of traits estimated using the mouse adult brain ST data (a) or the mouse embryonic brain ST data (b).

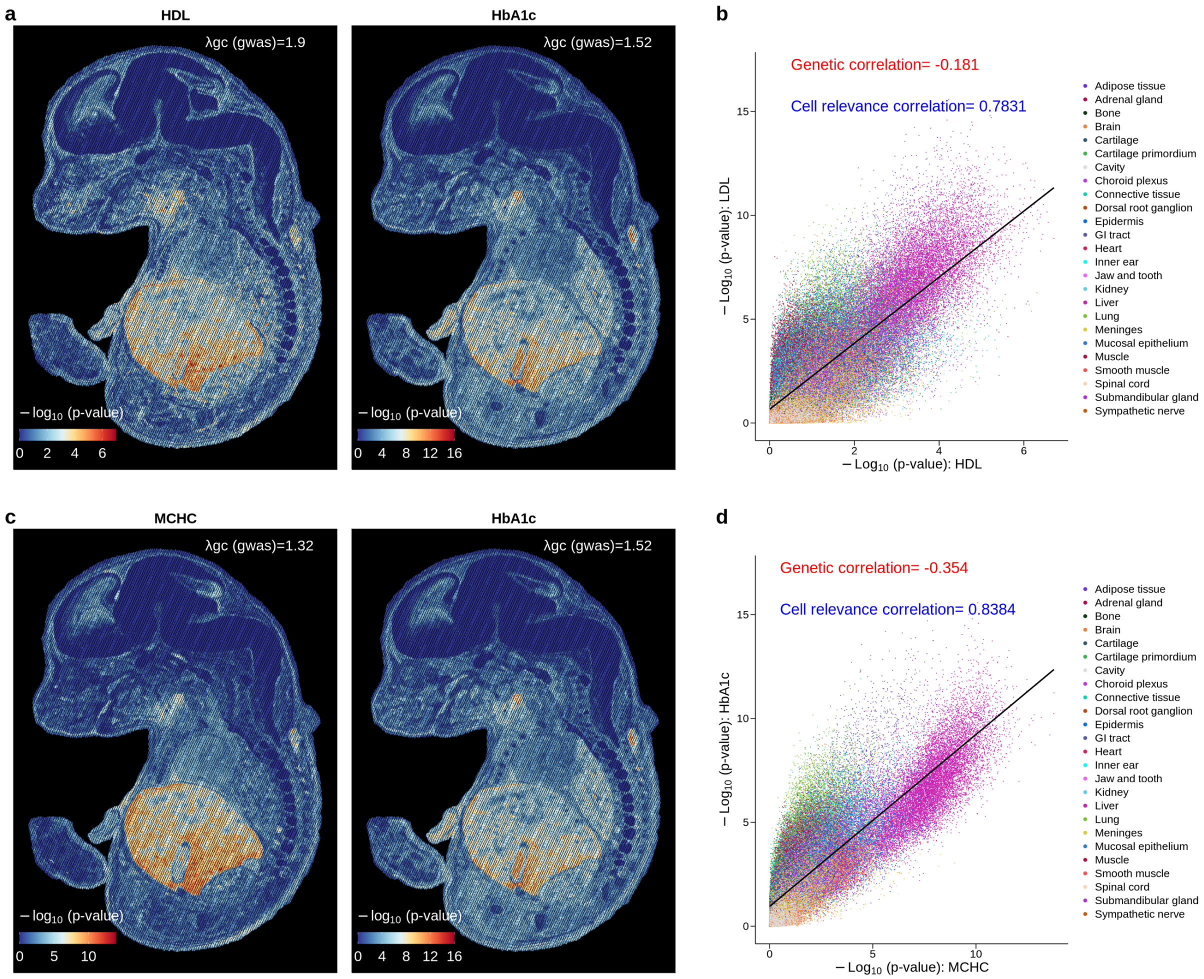


**Supplementary Figure 14. CRC vs. genetic correlation.** (a) gsMap results for HDL and HbA1c. Each point represents an individual spot, colored by the significance of its association with a trait. (b) CRC between HDL (x-axis) and HbA1c (y-axis), where each point represents an individual spot colored by its tissue type. (c) gsMap results for MCHC and HbA1c. (d) CRC between MCHC (x-axis) and HbA1c (y-axis), where each point represents an individual spot colored by its tissue type. The black line is the regression line.

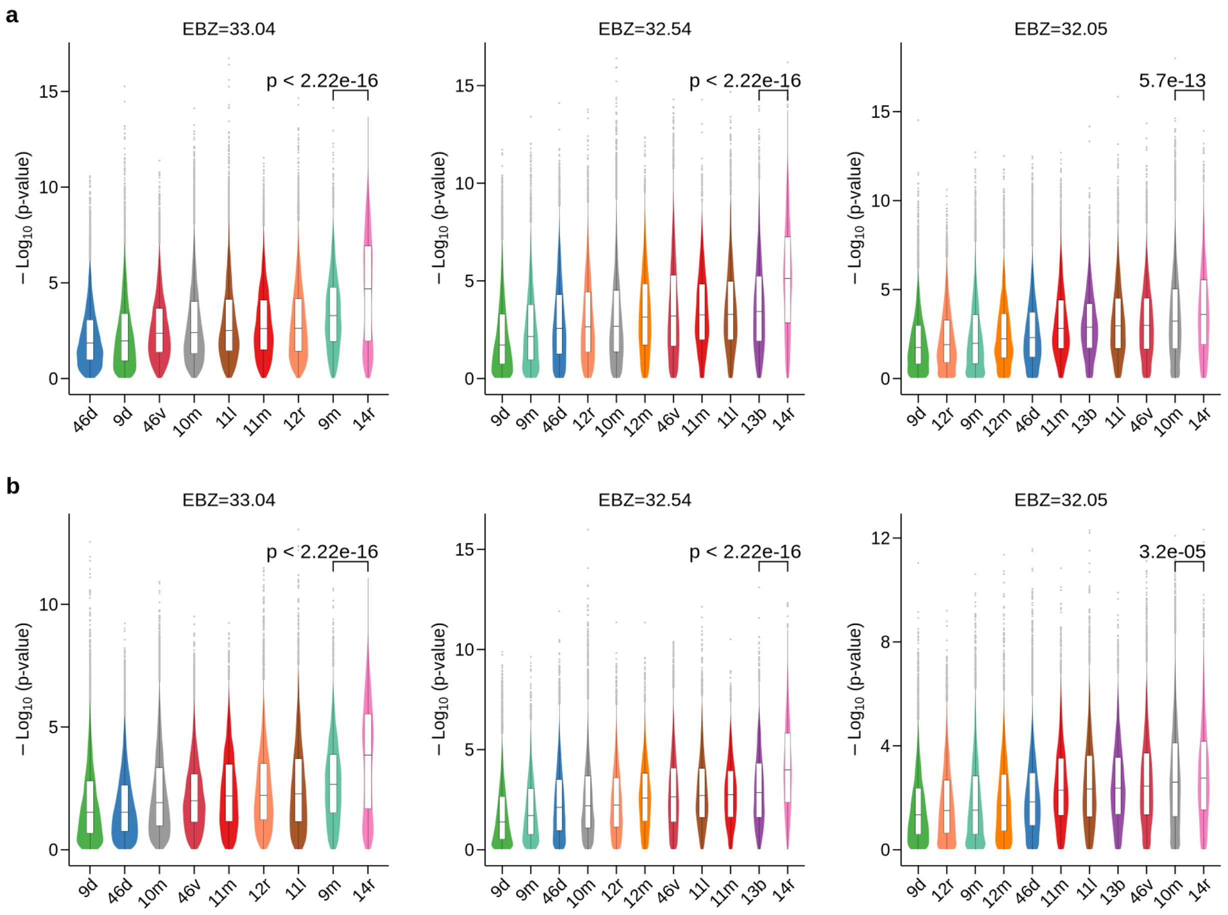


**Supplementary Figure 15. gsMap results in the PFC 14r region vs. the other PFC regions.** Violin plots of gsMap results in the PFC 14r region vs. other PFC regions for depression (a) and MDD (b). In each box, the central line denotes the median, notches represent the 95% CI, the box indicates the IQR, and whiskers extend up to 1.5 times the IQR, with outliers shown as individual dots.

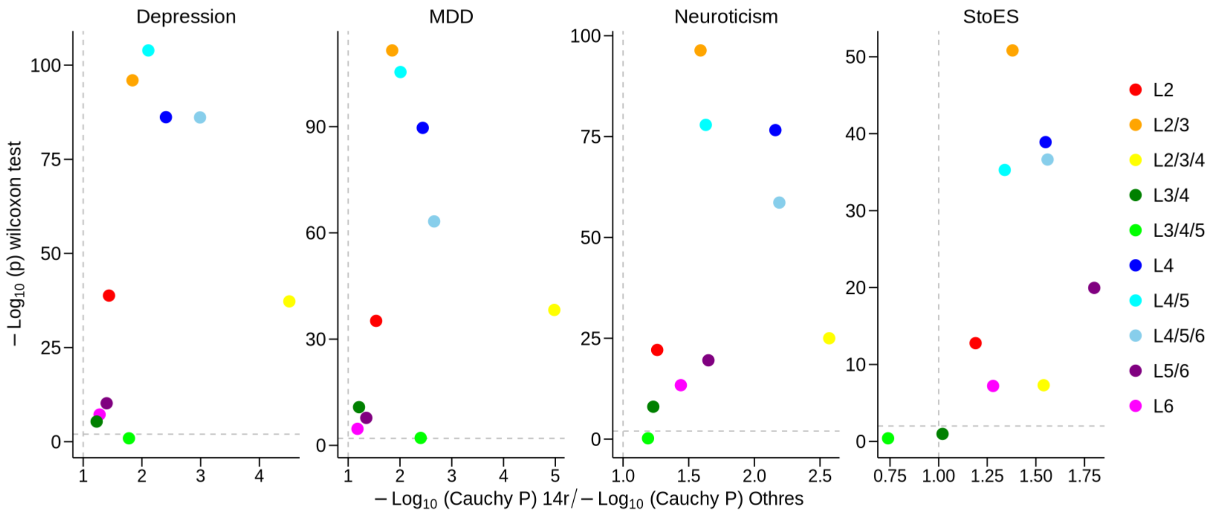


**Supplementary Figure 16. Comparison of gsMap results between the PFC 14r region and the other PFC regions for different glu-neuron subtypes.** The x-axis displays a ratio of -log10(*P* value) for association of a trait with a subtype of glu-neurons in the PFC14 region to that in all the other PFC regions, and the y-axis shows the Wilcoxon rank sum test *P* value for the difference between the two. Each data point represents a glu-neuron subtype. The vertical dashed line indicates the ratio of 1, and the horizontal dashed line indicates the significance threshold at FDR=0.01.

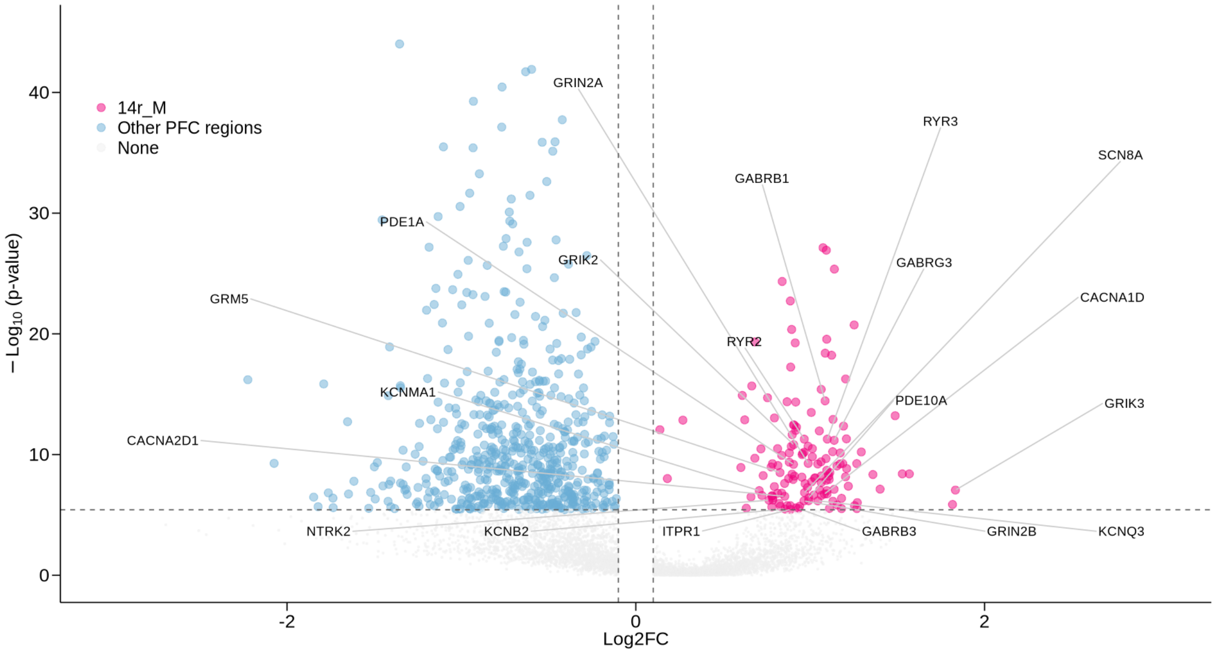


**Supplementary Figure 17. Genes highly expressed in the medial side of macaque PFC 14r.** The x-axis displays the log2 fold change (Log2FC) of gene expression in the medial side of PFC 14r compared to that in the other PFC regions; the y-axis displays the Wilcoxon rank sum test *P* value for the difference in gene expression. Each data point represents one gene. The vertical dashed lines indicate the Log2FC at ±0.1, and the horizontal dashed line indicates the significant threshold at FDR=0.05. The text shows the 20 psychiatric drug target genes overlapped with genes highly expressed in the medial side of PFC 14r.

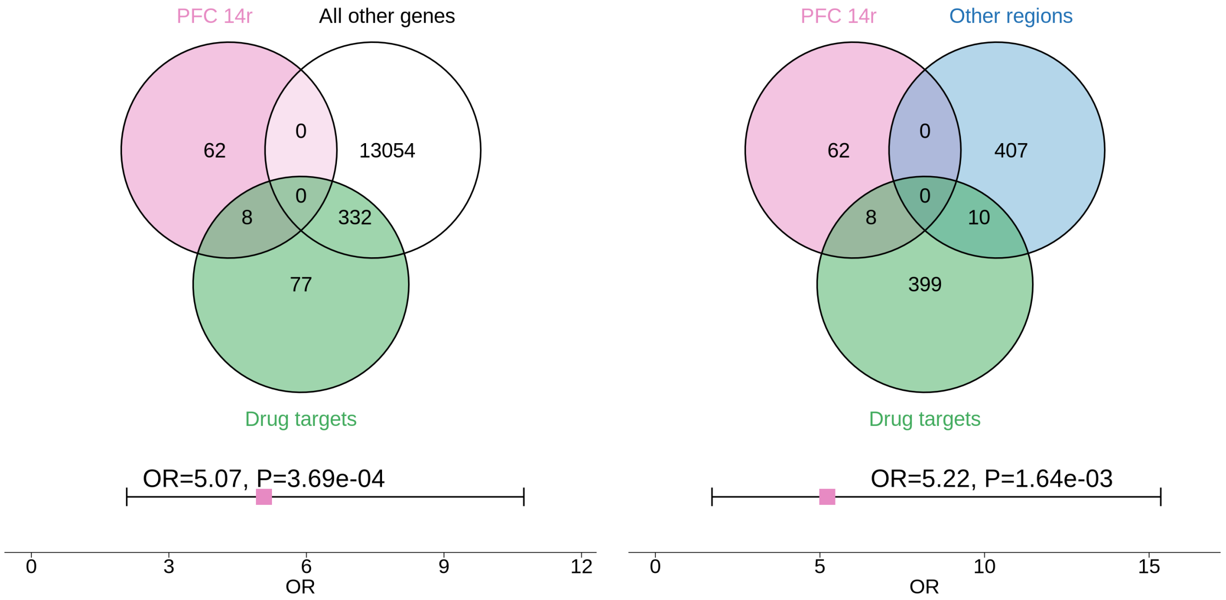


**Supplementary Figure 18. Enrichment of genes highly expressed in the PFC 14r region among psychiatric drug target genes.** This figure shows the fold-enrichment of genes highly expressed (at FDR<0.05) in the PFC 14r region among psychiatric drug target genes, when compared with all other detected genes (Left) or highly expressed genes in any of the other PFC regions (Right). The pink square denotes the OR, with the error bar representing the 95% CI.

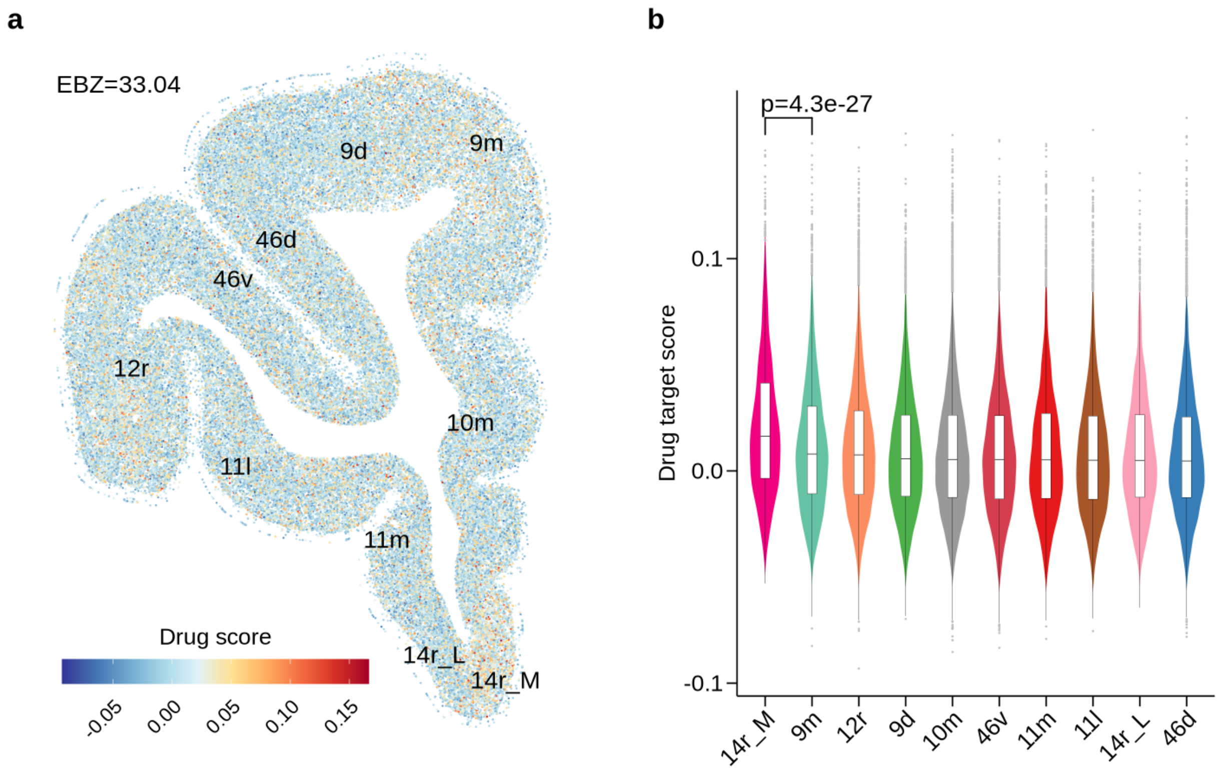


**Supplementary Figure 19. Psychiatric drug module score.** (a) Drug module score of 417 psychiatric drug target genes in the macaque PFC ST data, where each point represents a cell colored by its drug module score. (b) Violin plot of drug module score in the medial side of PFC 14r, compared to that in each of the other regions. The drug module score was estimated using the 'AddModuleScore' function in the Seurat[5 6](V4.4.0) R package with default settings. The black text shows the Wilcoxon rank sum test *P* value for the difference. In each box, the central line denotes the median, notches represent the 95% CI, the box indicates the IQR, and whiskers extend up to 1.5 times the IQR, with outliers shown as individual dots. 14r_L, 14r lateral side; 14r_M, 14r medial side.

### Supplementary Tables

We present the titles of supplementary tables here, and the full content of each table can be found in the corresponding Excel file.

#### Supplementary Table 1

GWAS summary statistics used in this study.

#### Supplementary Table 2

Macaque cortex ST sections.

#### Supplementary Table 3

Abbreviation of macaque cortex regions.

#### Supplementary Table 4

Identified drugs with target genes enriched in genes highly expressed in the medial side of PFC 14r.
